# Quantifying Geographic Heterogeneity in TB Incidence and the Potential Impact of Geographically Targeted Interventions in South and North City Corporations of Dhaka, Bangladesh: A model-based Study

**DOI:** 10.1101/2020.10.22.20217620

**Authors:** Sourya Shrestha, Mehdi Reja, Isabella Gomes, Yeonsoo Baik, Jeffrey Pennington, Shamiul Islam, Abu Jamil Faisel, Oscar Cordon, Tapash Roy, Pedro G. Suarez, Hamidah Hussain, David Dowdy

## Abstract

**Background:** In rapidly growing, high burden urban centers, identifying tuberculosis (TB) transmission hotspots and understanding potential impact of interventions can inform future control and prevention strategies.

**Methods:** Using data on local demography, TB reports, and patient reporting patterns in Dhaka South City Corporation (DSCC) and Dhaka North City Corporation (DNCC), Bangladesh, between 2010 and 2017, we developed maps of TB reporting rates across wards in DSCC and DNCC and identified wards with high rates of reported TB (i.e. “hotspots”) in DSCC and DNCC. We developed ward-level transmission models and estimated the potential epidemiological impact of three TB interventions: active case finding (ACF), mass preventive therapy (PT), and a combination of ACF and PT, implemented either citywide or targeted to high-incidence hotspots.

**Results:** There was substantial geographic heterogeneity in the estimated TB incidence in both DSCC and DNCC: incidence in the highest-incidence wards was over ten times higher than in the lowest-incidence wards in each city corporation. ACF, PT and combined ACF plus PT delivered to 10% of the population reduced TB incidence by a projected 7-9%, 13-15%, and 19-23% over five years, respectively. Targeting TB hotspots increased the projected reduction in TB incidence achieved by each intervention 1.4- to 1.8-fold.

**Conclusions:** The geographical pattern of TB notifications suggests high levels of ongoing TB transmission in DSCC and DNCC, with substantial heterogeneity at the ward level. Interventions that reduce transmission are likely to be highly effective, and incorporating notification data at the local level can further improve intervention efficiency.

## Introduction

Tuberculosis (TB) is the leading single-agent infectious cause of morbidity and mortality, with an estimated 10.0 million new TB cases and 1.5 million deaths worldwide in 2018.^1^ Despite the availability of effective treatment, TB incidence has not declined substantially in many high---burden countries, including Bangladesh, where an estimated 357,000 people developed new TB disease in 2018.^1, 2^ The End TB Strategy, launched by the World Health Organization as part of its post---2015 agenda, set goals to reduce TB incidence by 50% by 2025, and by 90% by 2035.^3^ Unfortunately, given the present slow decline in TB incidence of 1.5% per year, it is unlikely that these targets will be met unless concerted efforts are made to rapidly increase the rate of decline in TB incidence.

In high burden settings, a substantial proportion of incident TB occurs as a result of recent transmission.^4,5^ This is particularly true in densely populated urban centers, such as Dhaka, which have higher social contact rates, facilitated by factors such as use of mass public transportation, presence of slums and markets, and high rates of internal migration.^6-10^ Furthermore, it is known that TB, along with many of its common risk factors, such as low socioeconomic status,^11^ poor living conditions (e.g. crowding, poor ventilation in housing),^12-14^ migration status,^8,10^ and HIV infection,^15,16^ tends to cluster in hyperendemic “hotspots”. These high---incidence areas can act as reservoirs of infection and drive secondary transmission within the larger community.^12,17,18^

As such, interventions aimed at reducing transmission maybe critical to bring about decline in TB incidence in these settings.^19,20^ Furthermore, targeting hotspots may be more effective in reducing TB incidence at the local (e.g., city) level compared with interventions that are delivered to the general population without any attempt to prioritize those at highest risk.^21^ Therefore, in this study we aimed to understand the population level impact of TB interventions aimed at reducing TB transmission, and the added value of targeting hotspots with these interventions. Using data on TB notification, patient reporting patterns, and transmission models of TB, we estimate the impact of targeting potential TB interventions, namely active case finding (ACF, designed to reduce transmission by finding cases earlier) and preventive therapy (PT, designed to prevent reactivation of remote infection), to high-incidence geographical hotspots in Dhaka, Bangladesh.

## Methods

### Ward-level TB notification maps

We collected and aggregated notification data from TB reporting centers within each ward of Dhaka South City Corporation (DSCC) and Dhaka North City Corporation (DNCC) between the years 2010 and 2017— as of 2017, DSCC and DNCC consisted of 54 and 36 wards, respectively. Reporting centers provide TB treatment via Bangladesh’s Directly Observed Therapy, Short-Course (DOTS) program. We generated annual estimates of ward---level TB notification rates, calculated as the number of reported TB cases within each ward divided by the population of the ward (estimated using the 2011 national census with 5% annual growth rates). Using GIS data of the administrative boundaries of DSCC and DNCC, we then generated maps of the distribution of estimated TB notification rates in each ward.

To account for discrepancies between ward of TB notification (where patients were diagnosed) and ward of residence (where patients lived), we collected individual-level data on 2,980 patients diagnosed with TB from selected reporting centers in five DSCC wards and 532 patients in a single DNCC ward between 2017 and 2018. Using these data as a guide, we adjusted previously generated ward---level TB notification rates to reflect the observed distribution of TB cases notified in a given ward that comprised patients living in the ward of notification, patients living in adjacent wards (distributed equally across all adjacent wards), and patients living in non-adjacent wards (distributed equally across all other wards in the corresponding city corporation).

### Transmission Model

Drawing on our previous work,^12,17,19^ we constructed ward---specific epidemiological models to characterize transmission patterns and natural history of TB in all wards of DSCC and DNCC. Following a deterministic, compartmental model structure (Figure 1), each ward’s population was stratified into three compartments: TB uninfected, latent TB infection (LTBI, including post---treatment), and active (infectious) TB disease. We assumed that uninfected individuals, upon being infected with TB, progress either to LTBI or to active TB disease (“primary progression”). Latently infected populations could develop active TB disease either via reactivation or via reinfection followed by primary progression. We assumed that prior TB infection provides partial protection against future TB infection. Finally, we modeled diagnosis and successful treatment of TB disease as a return to the LTBI compartment. The model did not consider age-structure, drug resistance, or other risk factors that may affect TB natural history.

**FIGURE 1:**
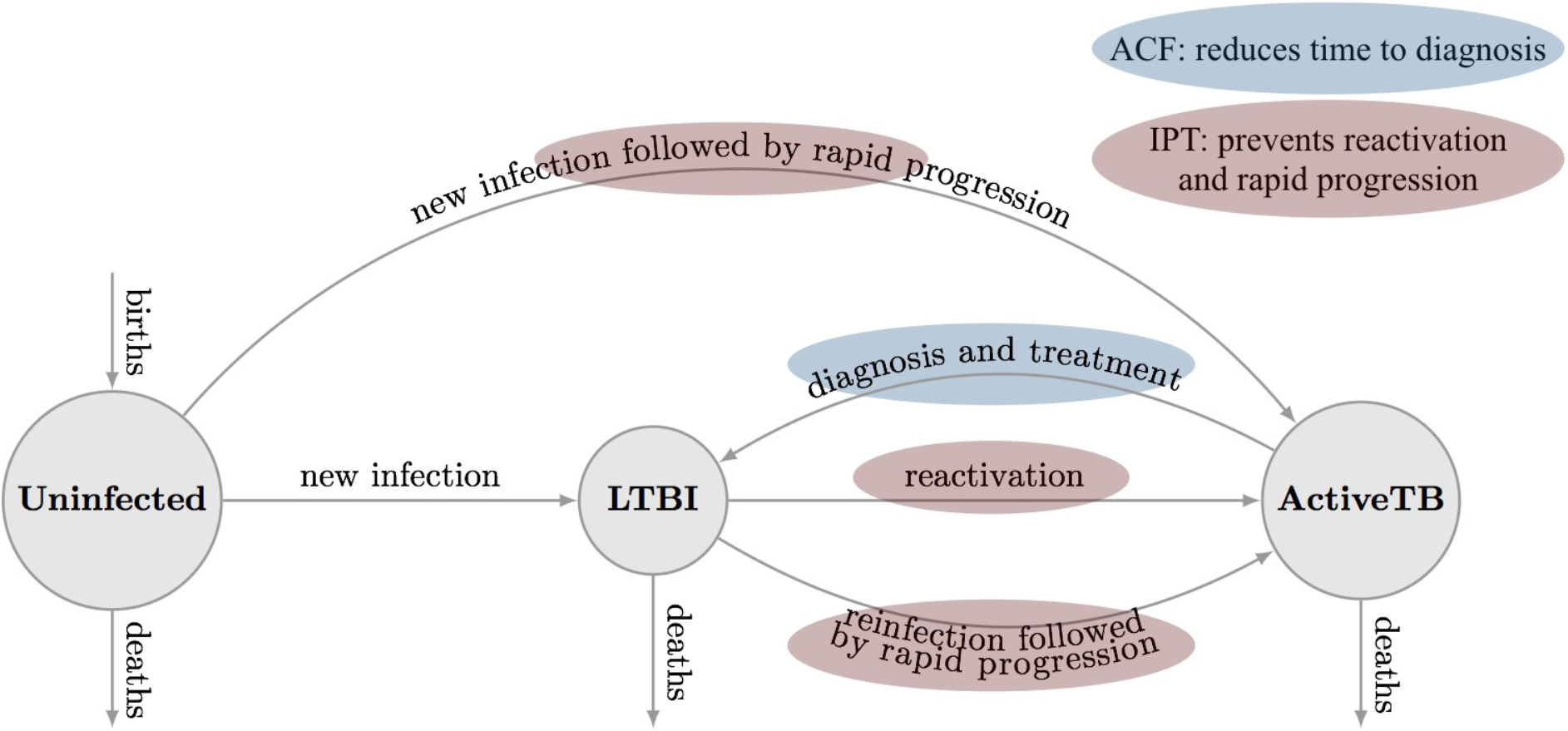
Schematic of Transmission Model. In this ward---specific compartmental model, the population was divided into three compartments based on their TB status: Uninfected (i.e. individuals who have not been exposed to TB), LTBI (i.e. individuals with latent TB infection), and Active TB (i.e. individuals with infectious TB disease). We modeled two interventions: Active Case Finding (ACF), which was modeled to reduce time to diagnosis and thus, resulting in an increase in the rate marked in blue; and Preventive therapy (PT), which was modeled to prevent reactivation and progression of disease and thus, resulting in a reduction in the rates marked in red.

We calibrated the models to ward---specific TB prevalence, based on the maps generated (as described above). Following WHO estimates, we assumed that 67% of incident TB cases in Bangladesh are reported. Other model parameters were taken from the published literature (See Table A1, Appendix II for details).^19^ To enable a simple, transparent model calibration process, we assumed that there were no significant secular trends in TB incidence at baseline.

### Model Scenarios

Genomic data (e.g., population-wide whole-genome sequencing) to inform the amount of ongoing TB transmission are not available for Dhaka or similar high-burden urban settings.^22^ As such, we modeled three different scenarios, each reflecting different levels of TB transmission at the ward level and each independently calibrated to the estimated TB incidence in Dhaka. These scenarios were chosen to reflect reasonable levels of transmission that could each be consistent with the observed epidemiology of TB in Dhaka.

1. **Low transmission**. In this scenario, an estimated 71% (interquartile range: 62%---79%) of incident cases in DSCC and 56% (interquartile range: 52%---61%) of incident cases in DNCC were due to recent transmission rather than reactivation of remote infection – a figure that is lower than estimated in models of Rio de Janeiro, Brazil,^12^ and other urban settings in countries where TB incidence is substantially lower than that of Bangladesh.
2. **Moderate transmission**. In this scenario, we used our *best a priori* estimates of TB transmission, as described in our model of TB transmission in Karachi, Pakistan.^19^ This resulted in 82% (interquartile range: 76%---87%) of incident cases in DSCC and 70% (interquartile range: 66%---75%) of incident cases in DNCC being due to recent transmission. This scenario is used as the reference for all results presented below.
3. **High transmission**. Here, we assumed higher transmission rates, similar to the “high-transmission” scenario in our model of Karachi,^19^ such that the vast majority of TB is due to recent transmission. This resulted in 88% (interquartile range: 83%---91%) of incident cases in DSCC and 79% (interquartile range: 75%---83%) of incident cases in DNCC being due to recent transmission events.

### Interventions

We modeled three different TB interventions; (i) active case finding (ACF); (ii) mass preventive therapy (PT); and (iii) ACF and PT combined, with each intervention achieving a population-level coverage of 10% in DSCC and DNCC separately. For ACF, we assumed that implementation would reduce time to diagnosis by one---third (i.e., 33.3% reduction in the average time to diagnosis). For PT, we assumed adherence levels of 60% and efficacy of 80% in reducing reactivation and rapid progression of infections that existed at the time of the intervention. For the combined intervention, we assumed that both interventions, ACF and PT, would be implemented in the same population with independent effects. For simplicity, we assumed rapid scale-up of each intervention to the target level specified.

For all three interventions, we modeled two implementation strategies, either a citywide implementation (in which 10% of the entire population of DSCC and DNCC received the intervention, regardless of ward of residence), or a targeted implementation (in which the interventions were targeted to high---incidence wards, but at a higher coverage such that the same number of people were covered as in the citywide implementation). For DSCC, we selected the 12 wards with the highest TB notification rates in 2017, which comprised approximately 20% of the total population of DSCC. Similarly for DNCC, we selected the 6 wards with highest TB notification rates between 2015 and 2017; these wards comprised approximately 20% of the total population of DNCC. We assumed that 50% of the population in these wards would be covered by each intervention under targeted implementation, thereby achieving a population-level coverage of 10% in each city corporation.

### Primary Outcome

The primary outcome was the projected reduction in TB incidence in DSCC and DNCC, five and ten years after implementation of each intervention, comparing targeted implementation of the intervention in high incidence “hotspots” versus untargeted citywide implementation.

### Sensitivity analyses

To explore the sensitivity of the model results to the changes in model parameters, we conducted multivariate uncertainty analyses. We generated 10,000 parameter sets for DSCC and DNCC separately using Latin Hypercube Sampling, carried out model simulations for each parameter set, and estimated partial rank correlation coefficients (PRCC), between the model parameters and key model outcomes, relative reduction in 10-year TB incidence achieved through ACF, PT and a combination of both via targeted implementation compared to a citywide implementation.^23^ (See Appendix II for details.)

## Results

### Spatiotemporal Patterns of TB in DSCC and DNCC

TB notification data during the seven-year period between 2010 and 2017 suggests that, while TB notification rates are generally higher in DSCC compared to DNCC, TB is highly heterogeneous at the ward level in both city corporations (Figures 2 and 3). Unadjusted ward-level notification rates in 2010 in both DSCC and DNCC ranged from 60.9 to 2,822.6 per 100,000 per year, over 40-fold difference.

**FIGURE 2:**
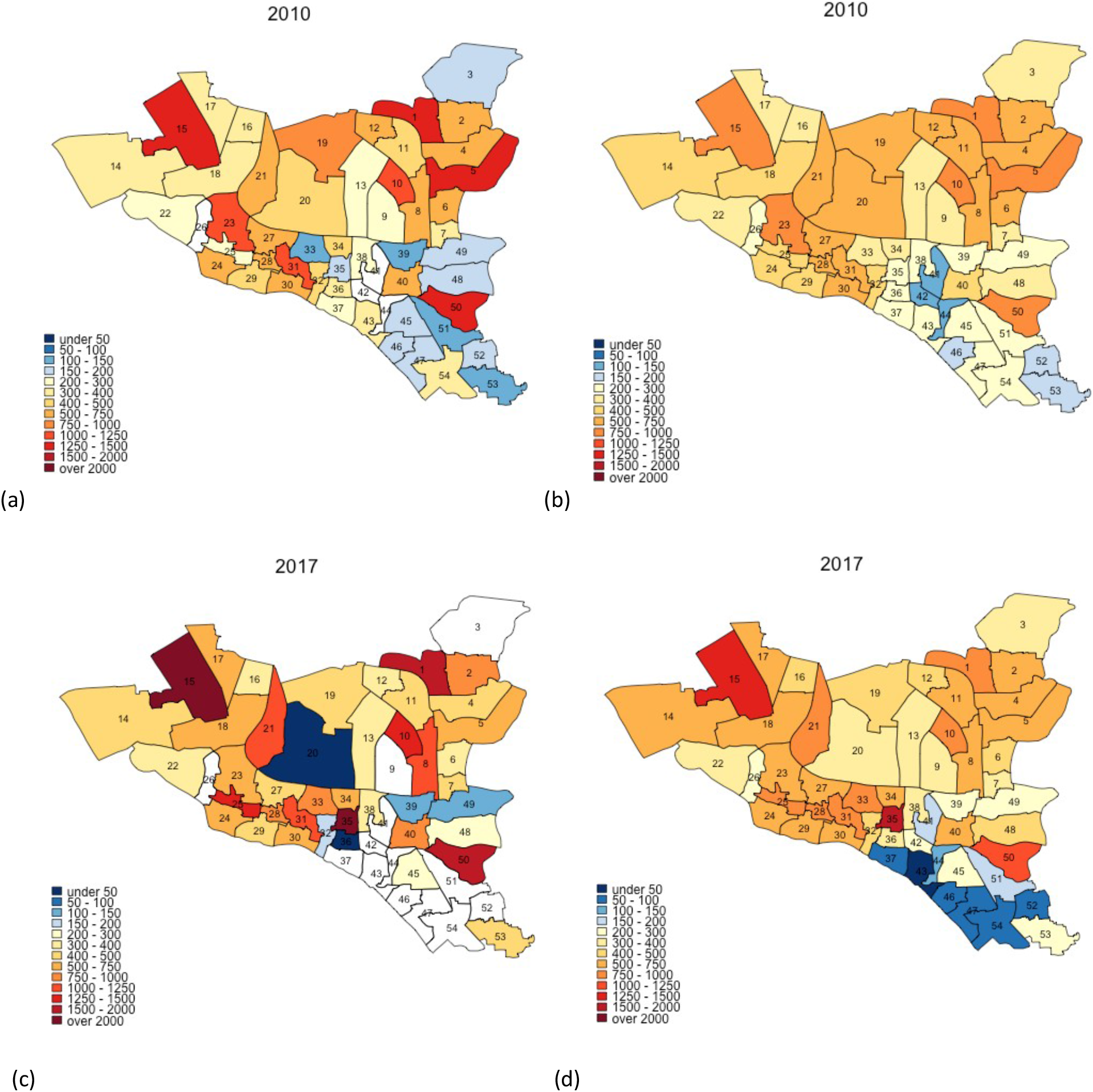
Maps of DSCC with estimated TB notification rates in 2010 and 2017. Panels (a) and (c) show unadjusted notification rates (in units per 100 000 per year), whereas panels (b) and (d) show corresponding notification rates after adjustment for observed correlations between ward of residence and ward of reporting center. Panels (a) and (b) show data from 2010, and panels (c) and (d) represent 2017 data.

**FIGURE 3:**
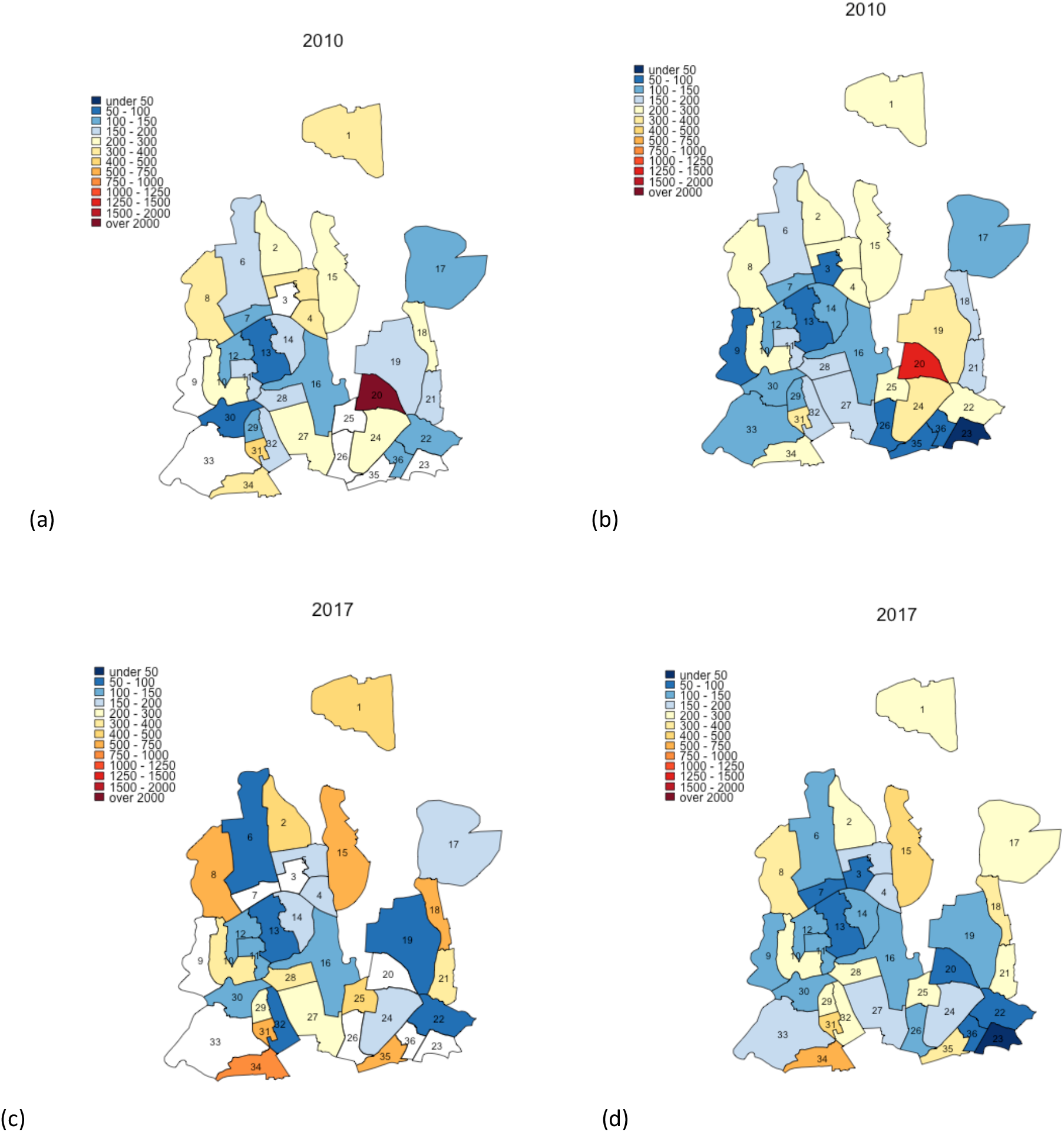
Maps of DNCC with estimated TB notification rates in 2010 and 2017. Panels (a) and (c) show unadjusted notification rates (in units per 100 000 per year), whereas panels (b) and (d) show corresponding notification rates after adjustment for observed correlations between ward of residence and ward of notification. Panels (a) and (b) show data from 2010, and panels (c) and (d) represent 2017 data.

Using individual-level data from selected reporting centers, we estimated that 18-50% of reported cases resided in the ward in which the reporting center was housed, 12-37% resided in neighboring wards (wards within the city corporation sharing boundaries with the reporting center), and the remaining resided elsewhere in Dhaka. For each ward, we therefore adjusted notification rates to more closely reflect potential patterns of patient residence by attributing 50% of the reported cases to the ward housing the reporting center, 40% to adjacent wards, and 10% equally to all wards in the city corporation. Even after adjusting for potential clustering of reporting in this fashion, substantial geographic heterogeneity in TB incidence persisted in both DSCC and DNCC; distinct “hotspots” with high TB notification rates and “cold” patches with low TB notifications were still observed. This pattern of geographic heterogeneity persisted and intensified over time, as depicted by TB notification maps of DSCC and DNCC, which show darker shades of red and blue in 2017 (Figures 2 and 3, panels c and d) than in 2010 (panels a and b).

Changes in TB notification rates between 2010 and 2017 were also heterogeneous across wards. For example, in DSCC, the southern and north---central sections had prominent declines in TB notification rates, whereas the central and north eastern parts experienced large increases (Figure 4b; blue patches indicate declines, and red and orange patches indicate increases). In DNCC, the central area had marked declines, whereas the eastern and southern areas experienced large increases (Fig 4d).

**FIGURE 4:**
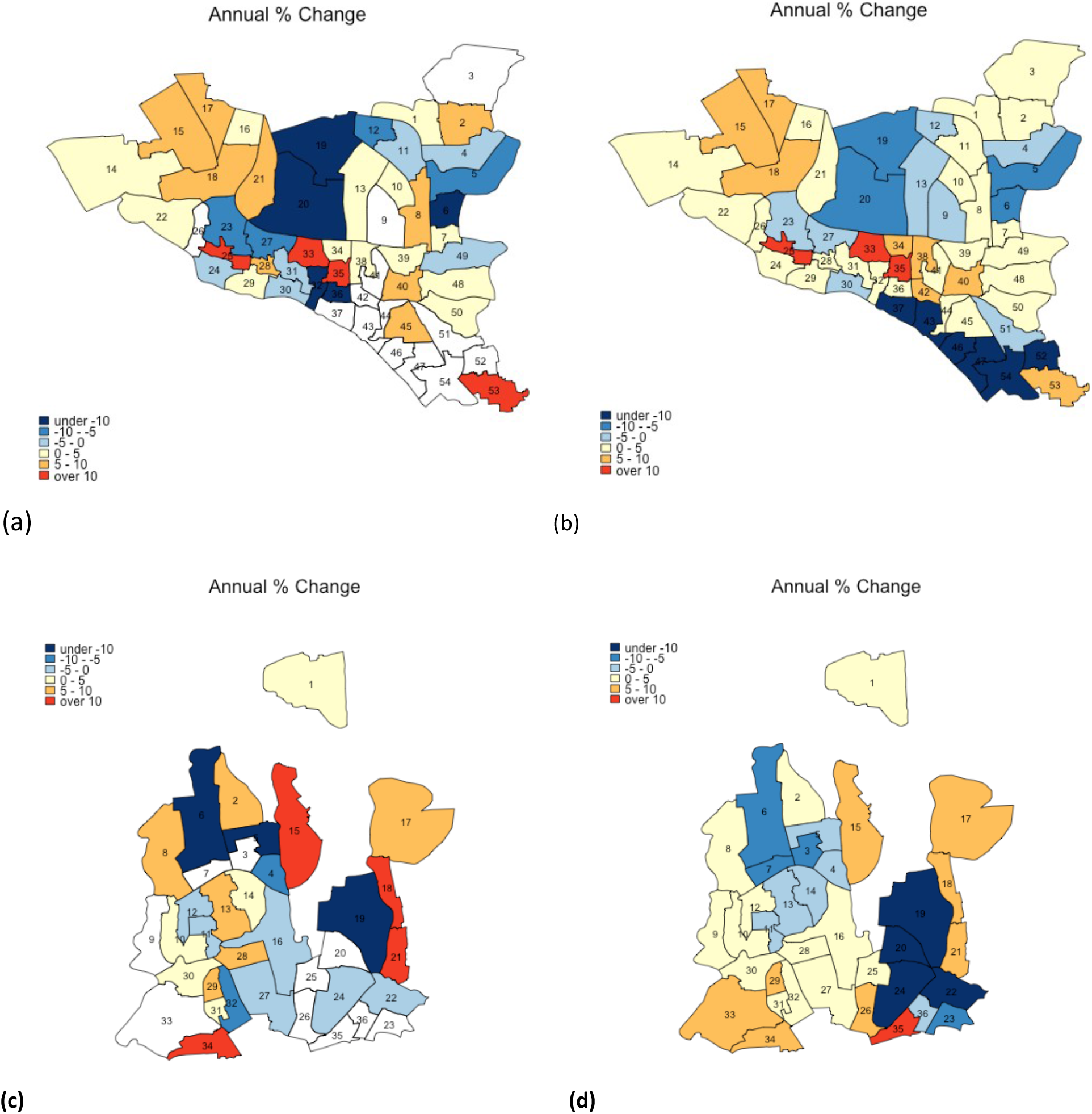
Annual percent change in TB incidence in DSCC and DNCC wards between 2010 and 2017. Panel (a) and panel (c) give the average annual changes (% per year) in estimated TB incidence at the ward level in DSCC and DNCC, respectively, between the years 2010 and 2017. Panel (b) and panel (d) show corresponding data after adjustment for correlations between ward of residence and ward of notification. Blue shading indicates a decline in TB incidence during the 7---year period (with darker shades representing steeper declines), whereas red shading indicates an increase (with darker shades representing greater increases).

### Epidemiological impact of TB interventions

Active case finding (ACF) implemented throughout DSCC over a five-year period and randomly targeting 10% of the population was projected to reduce TB incidence by 9.0%; when targeted to the 12 wards with the highest TB incidence, this impact grew to a 14.6% projected reduction (Figure 5). The corresponding impact of preventive therapy (PT) was a 15.2% (untargeted) and 22.3% (targeted) reduction in five-year incidence, and when ACF and PT were combined, the greatest reductions in five-year incidence were achieved: 22.6% if untargeted and 27.7% if targeted to the 12 highest-incidence wards (Figure 5).

**FIGURE 5:**
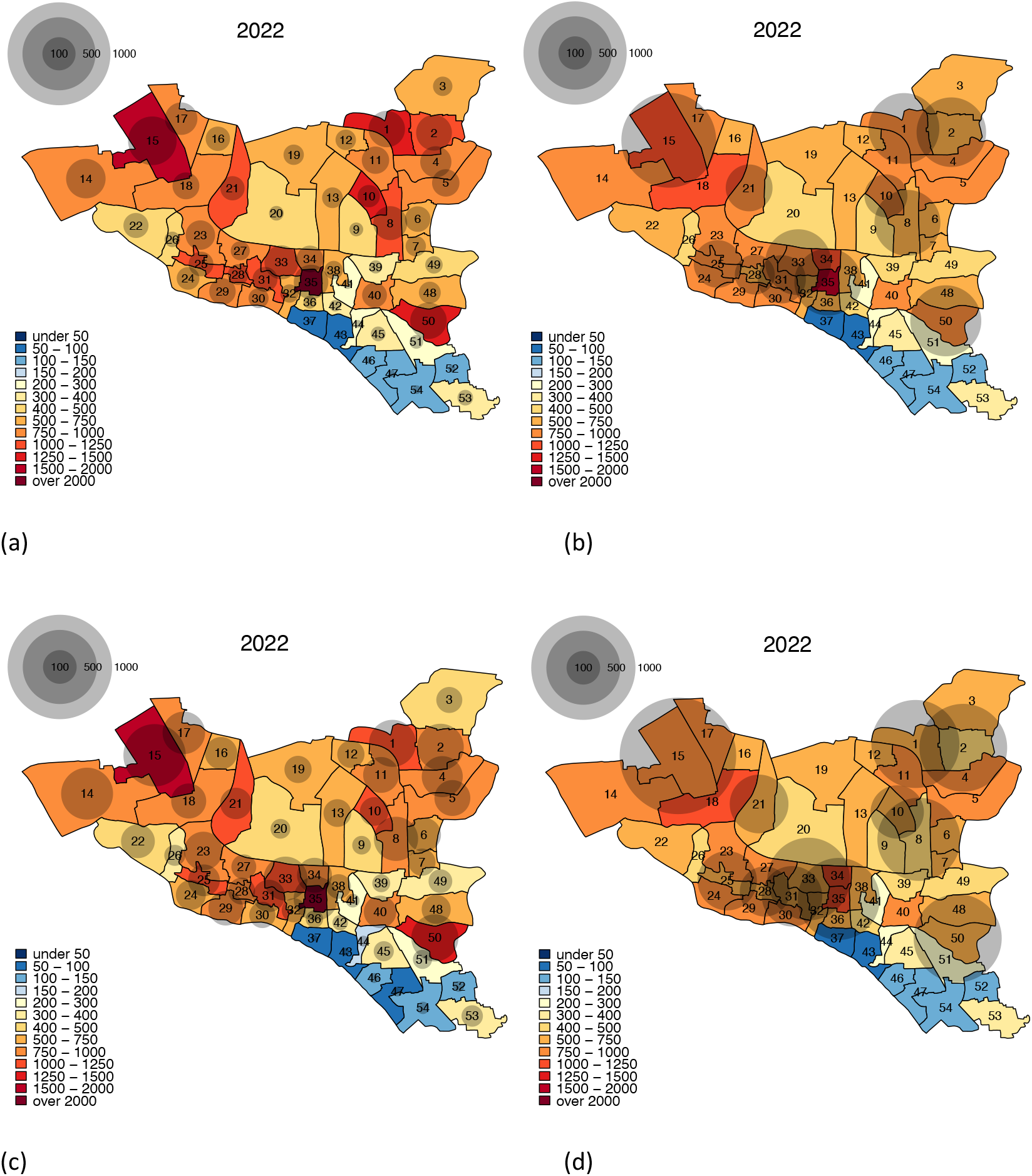
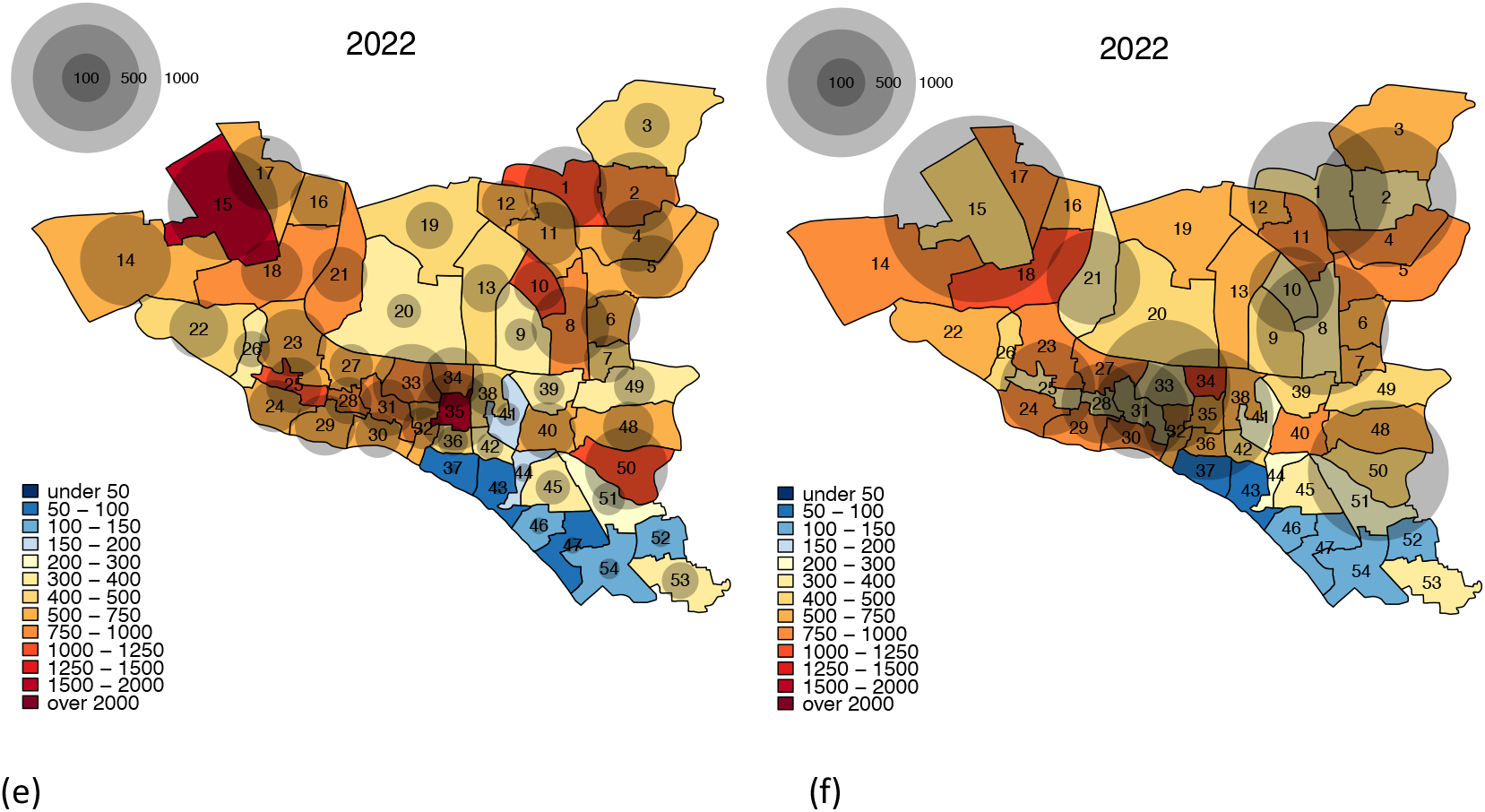
Impact of TB interventions on ward-level TB incidence in DSCC after five years. The colors for each ward depict the projected TB incidence after five years of intervention, and the bubbles indicate the absolute size of the reductions (the reduction in the number of incident TB cases achieved by the intervention in year 5). Panel (a) represents city---wide active case finding, (b) targeted case finding, (c) city---wide preventive therapy, (d) targeted preventive therapy, (e) combination of citywide active case finding and preventive therapy, and (f) combination of targeted active case finding and preventive therapy.

**FIGURE 7:**
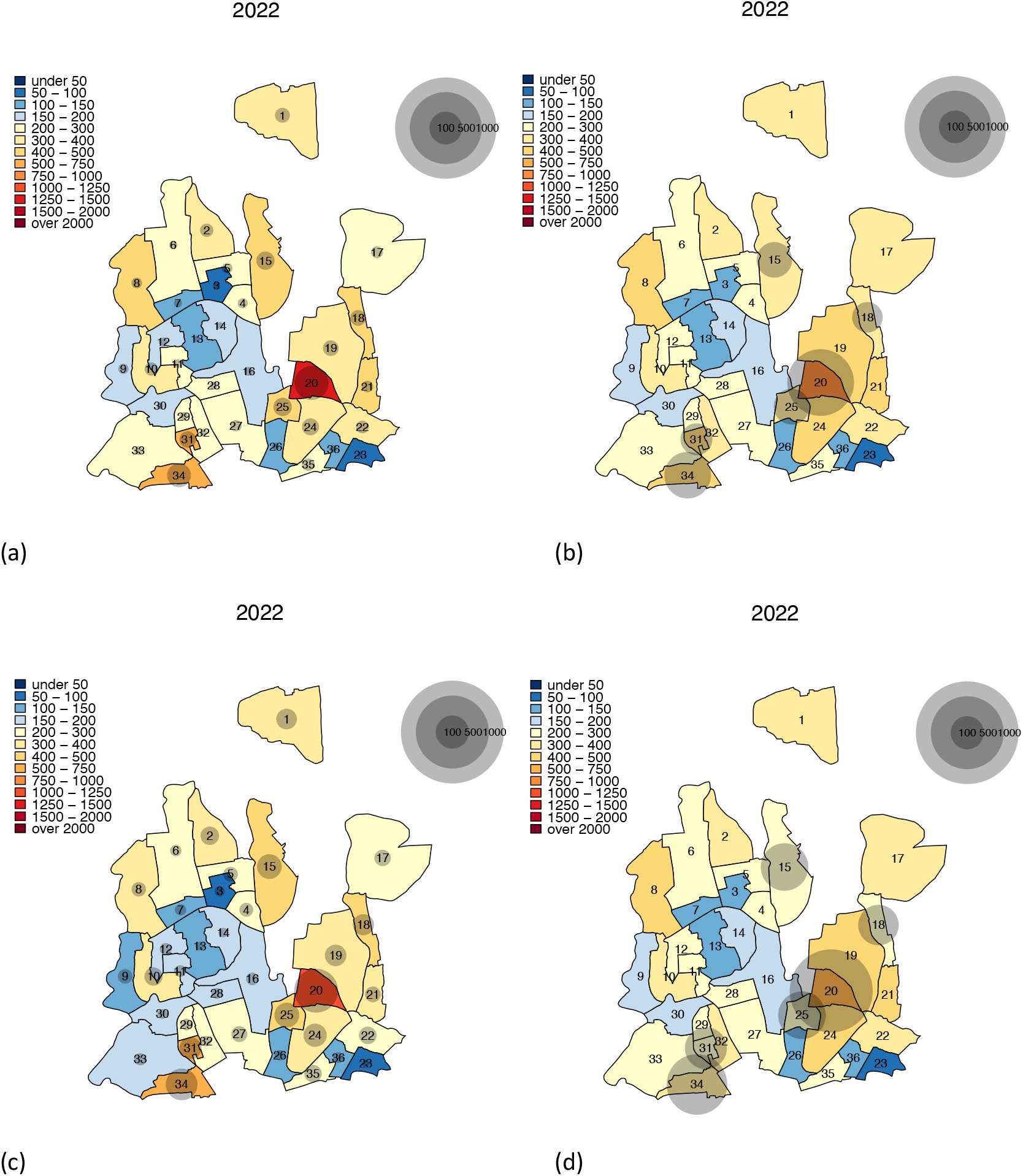
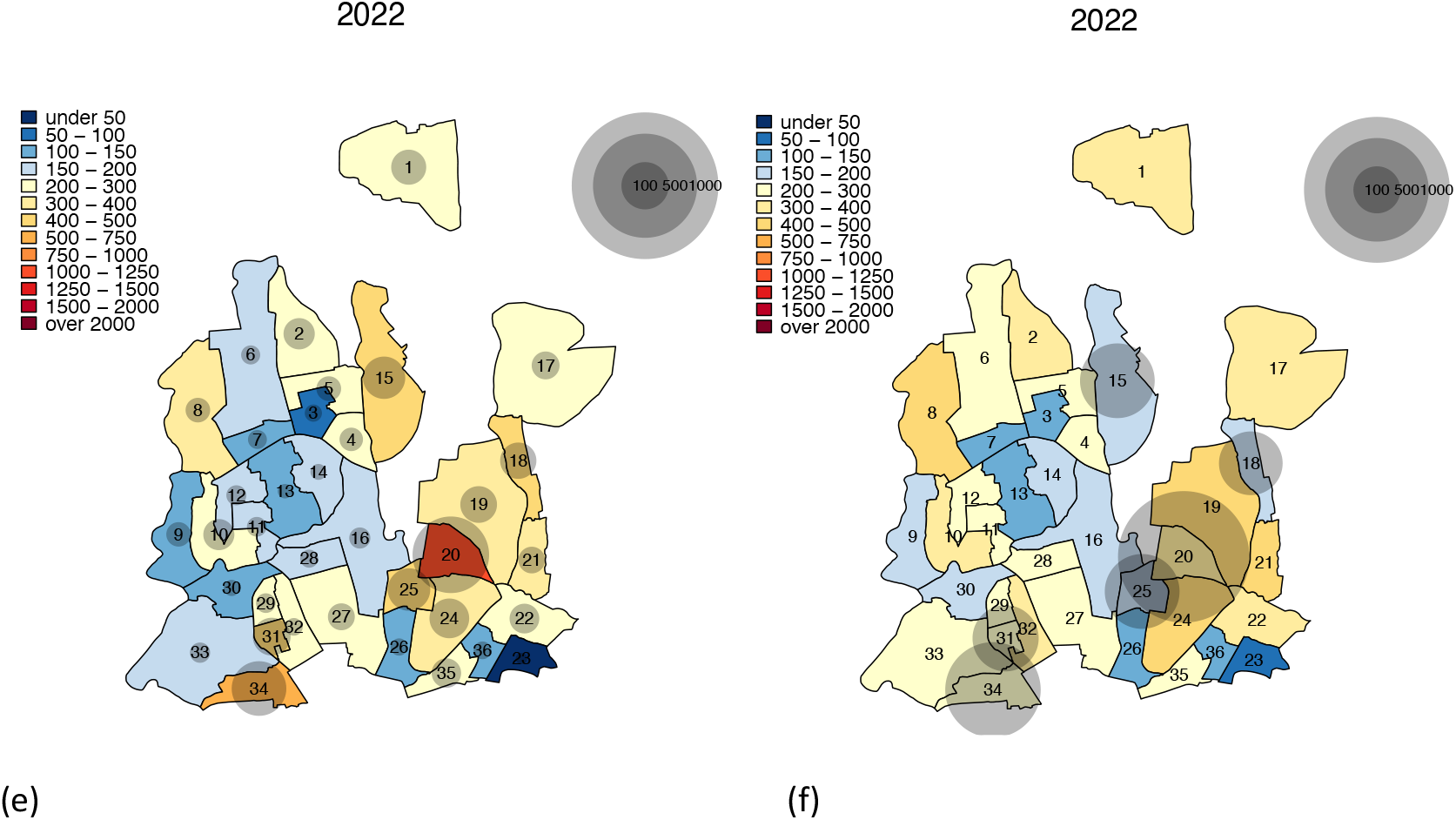
Impact of TB interventions on ward-level TB incidence in DNCC after five years. The colors for each ward show the projected TB incidence after five years of intervention, and the bubbles indicate the absolute size of the reductions (the reduction in the number of incident TB cases achieved by the intervention in year 5). Panel (a) represents city---wide active case finding, (b) targeted case finding, (c) city---wide preventive therapy, (d) targeted preventive therapy, (e) combination of citywide active case finding and preventive therapy, and (f) combination of targeted active case finding and preventive therapy.

The impact of TB interventions in DNCC was slightly lower than in DSCC, reflecting the lower burden of TB incidence and TB transmission in DNCC relative to DSCC (Figure 6). Projected reductions in five-year TB incidence in DNCC were: 7.0% from citywide ACF, 13.9% from hotspot-targeted ACF, 13.0% from citywide PT, 22.8% from targeted PT, 18.9% from citywide combined ACF and PT, and 28.2% from targeted ACF and PT. Over a ten-year time horizon, the projected epidemiological impact of all citywide interventions grew by an additional 18-41%; this grew to 19.8-44.8% at 20 years. Notably, the relative added benefit of targeting was greatest at earlier timepoints. For example, ACF deployed in a targeted fashion in DSCC was 1.6 times more impactful compared to citywide ACF at year 5, but only 1.4-times at year 20, suggesting that the relative value of targeting can wane over time. The relative added benefit of the targeting diminished when both interventions, ACF and PT were implemented in a targeted fashion. For example, at year 5 in DSCC, ACF and PT when applied separately, was respectively, 1.6 and 1.5 times impactful when targeted. However, when combined, the relative impact of targeting was only 1.2 times more. The impact of interventions also increased with the intensity of transmission, with the high---transmission scenario leading to estimates of impact at least 20% greater than those in the low---transmission scenario for all interventions. However, the relative benefit of geographically targeted implementation also decreased with higher levels of transmission; for example, targeted implementation of ACF in DNCC was estimated to generate a reduction in TB incidence that was 2.0 times greater than untargeted implementation in the low transmission scenario, compared to 1.7 times in the high transmission scenario (Figures S5 and S6 in the Appendix I). Finally, the results from multivariate uncertainty analyses show that targeted interventions have greatest impact in settings where more incident TB is due to recent rather than remote infection. The model parameters that correlated most strongly with the relative value of targeting interventions were the level of protection against reinfection, the rate of TB diagnosis, and the rate of rapid progression: increase in any one of these increases the proportion of incident TB that are due to recent infection compared to remote (See Appendix II).

## Discussion

In this study, we aimed to assess the benefits of potential TB interventions, specifically active case finding (ACF) and mass preventive therapy (PT), in Dhaka, Bangladesh – a high-incidence, densely urban South Asian city. We found that TB is geographically heterogeneous across wards, with ward-level notification rates varying by more than a factor of ten. Interventions in Dhaka to actively find TB cases and to prevent reactivation disease have the potential to effect substantial and rapid declines in TB incidence. For example, covering 10% of the population with active case finding and preventive therapy could reduce TB incidence in Dhaka by about 20% within five years. Targeting these interventions to the wards with highest TB notification rates could magnify the impact of these interventions still further, such that nearly 30% reductions in TB incidence could be achievable within five years. These results may help motivate the implementation of interventions to reduce TB transmission in South Asian megacities, and to collect data at the district level that could help inform evidence-based targeting of those interventions to high-incidence hotspots.

Geographic heterogeneity is a hallmark of most infectious diseases, including vector-borne diseases, such as malaria and dengue virus,^24-26^ and sexually transmitted diseases, such as gonorrhea, chlamydia and syphilis.^27^ For many of these infections, it has been recommended that interventions be targeted to high-incidence hotspots. Nevertheless, TB differs from most other infectious diseases, particularly in terms of its airborne route of infection and lengthy/highly variable trajectory of latency and disease, which may mitigate the degree of geographic heterogeneity and the impact of such heterogeneity on disease transmission. Understanding the dynamics of geographic heterogeneity in TB incidence can therefore not only inform the prioritization of existing TB interventions and resources, but can also add insight into the natural history and transmission patterns of *M. tuberculosis*, the most deadly human pathogen.^28-30^ Such investigations may be particularly useful in urban settings such as Dhaka, where TB transmission is particularly intense and notification data are available at the scale of small administrative units.

These results lay the groundwork for future modeling analyses, including a more detailed characterization of patient reporting patterns and mixing rates, as well as the integration of demographic, socio-economic, and TB care-seeking factors. Incorporation of genomic data could also refine our interpretation of TB incidence.^22^ Specifically, since the projected impact of TB interventions depends on the degree to which incident TB disease reflects recent transmission versus reactivation, more accurate estimates of the proportion and geographic distribution of new cases due to recent transmission can help refine estimates of intervention impact. Such additional analyses can also better quantify heterogeneity and help validate findings from simpler models such as the one presented here.

In addition to informing city-level policy (in collaboration with partners such as the Bangladesh National Tuberculosis Program, which contributed to this work), our model findings can also help motivate the collection of finer-resolution data on TB notifications in Dhaka and other similar settings. Our results illustrate how the ability to accurately identify high-incidence hotspots and assess reporting and mixing patterns, at an appropriate and actionable spatial scale (such as the ward level in Dhaka) can help to harness the full potential of geographically targeted TB interventions. In assessing the relative benefit of targeting actual interventions (rather than the stylized interventions presented here), feasibility and cost-effectiveness of delivering interventions at the local scale must also be considered.

As with any modeling analysis, these findings should be interpreted in light of certain limitations. For example, empirical data were not available to inform certain important considerations such as the movement of individuals between wards and city corporations. This forced us to adopt a number of simplifying assumptions, such as stability in the relative size of ward populations, which could affect our results in two distinct ways. First, discrepancies between patients’ place of residence and place of notification could affect our ability to accurately assess the geographic distribution of TB risk and incidence from notification data alone. Because TB transmission largely occurs within households and communities, tracking patients by their place of residence may be a more accurate measure of capturing the spread of TB than place of presentation.^19^ In our study, we partially accounted for these discrepancies through adjustment based on patient-level data, which we collected from reporting centers in five DSCC wards and one DNCC ward. In this analysis, we found that nearly half of all cases were reported in wards where patients did not live. These findings suggest that a systematic assessment of reporting patterns throughout DSCC and DNCC is necessary to more comprehensively address this concern. Second, movement of individuals between wards can further drive the spread of TB within the city; for example, high-incidence hotspots can fuel TB in many other parts of the city if there is a large amount of movement between the hotspot and the other parts of the city. ^10,12^ While such mixing is generally difficult to quantify for an airborne disease, a lack of data on between-ward mobility may result in an underestimation of the impact of targeted interventions. Geographically targeted interventions have been shown to have higher relative benefit when there is more mixing between individuals in the hotspots and the general population.^12,17^ Given the importance of mixing, there is a need to better understand the mobility and migration of high-risk populations (e.g. commuting patterns), from the perspective of airborne transmission events.

In summary, this mathematical model of TB transmission in DSCC and DNCC suggests that both active case finding and preventive therapy can achieve important reductions in TB incidence over a five-year period. If these interventions are combined and targeted to those wards with the highest TB notification rates (as identified using ward-level data), the achievable reduction in incidence can approach 30% within five years. The success of these interventions is only possible if carried out in conjunction with strengthening of existing diagnostic and treatment services, which would allow for appropriate diagnosis and treatment of an expanded number of individuals, without a loss in quality. These findings support efforts to intensify active TB case finding and preventive therapy in Dhaka, strengthen existing TB diagnostic and treatment services, and collect additional supporting data to further tailor the implementation of these interventions to those populations most affected by high rates of TB transmission.

## Data Availability

Data used in the manuscript will be made available.

