## Supplementary materials for "Quantifying Geographic Heterogeneity in TB Incidence and the Potential Impact of Geographically Targeted Interventions in South and North City Corporations of Dhaka, Bangladesh: A model-based Study"

**APPENDIX 1. Supplementary Results**

TABLE S1: **Projected percentage reduction in TB incidence in DSCC after 10 years of TB interventions.**

| *Ward* | *ACF-citywide* | *ACF-targeted* | *PT-citywide* | *PT-targeted* | *Combination-citywide* | *Combination-targeted* |
|---|---|---|---|---|---|---|
| **44** | 7.3 (4.8-9.9) | | 13.5 (10.6-16.4) | | 19.4 (14.6-24.0) | |
| **54** | 6.7 (4.4-9.0) | | 12.7 (10.2-15.4) | | 18.1 (13.9-22.4) | |
| **53** | 8.8 (5.8-11.5) | | 15.1 (11.7-18.2) | | 22.1 (16.5-27.0) | |
| **52** | 6.8 (4.5-9.1) | | 12.8 (10.2-15.5) | | 18.3 (14.0-22.7) | |
| **47** | 6.6 (4.4-8.8) | | 12.6 (10.1-15.2) | | 17.9 (13.7-22.1) | |
| **46** | 6.7 (4.5-9.0) | | 12.8 (10.2-15.4) | | 18.1 (13.9-22.4) | |
| **12** | 11.0 (7.5-13.5) | | 17.6 (13.7-20.4) | | 26.1 (19.8-30.7) | |
| **43** | 6.4 (4.3-8.6) | | 12.4 (10.0-14.9) | | 17.5 (13.5-21.6) | |
| **45** | 8.8 (5.8-11.5) | | 15.2 (11.8-18.2) | | 22.2 (16.5-27.1) | |
| **51** | 7.8 (5.1-10.4) | | 14.1 (11.0-17.0) | | 20.3 (15.2-25.1) | |
| **37** | 6.5 (4.3-8.7) | | 12.6 (10.1-15.1) | | 17.8 (13.7-21.9) | |
| **50*** | 15.1 (12.1-17.0) | 50.9 (41.7-56.3) | 22.3 (18.9-24.2) | 69.9 (62.8-62.8) | 33.6 (28.2-36.8) | 81.6 (74.0-85.7) |
| **42** | 9.0 (5.9-11.7) | | 15.3 (11.8-18.4) | | 22.4 (16.7-27.4) | |
| **7** | 10.4 (7.1-13.1) | | 17.0 (13.3-20.0) | | 25.1 (19.1-30.0) | |
| **36** | 10.0 (6.7-12.7) | | 16.5 (12.7-19.5) | | 24.3 (18.2-29.2) | |
| **30** | 12.3 (8.9-14.8) | | 19.1 (15.3-21.9) | | 28.6 (22.4-33.0) | |
| **32** | 11.3 (7.9-13.9) | | 18.0 (14.2-20.9) | | 26.8 (20.5-31.4) | |
| **48** | 10.9 (7.6-13.5) | | 17.6 (13.8-20.5) | | 26.0 (19.9-30.8) | |
| **40** | 12.8 (9.4-15.2) | | 19.7 (15.9-22.3) | | 29.5 (23.3-33.7) | |
| **29** | 12.5 (9.0-14.9) | | 19.3 (15.4-21.9) | | 28.8 (22.6-33.1) | |
| **35*** | 16.9 (14.7-18.2) | 56.6 (49.9-60.5) | 24.2 (21.8-25.6) | 74.1 (69.1-69.1) | 36.9 (32.9-39.1) | 86.2 (81.0-89.1) |
| **28*** | 13.9 (10.6-16.0) | 47.0 (37.1-53.4) | 20.8 (17.2-23.2) | 66.9 (59.1-59.1) | 31.3 (25.4-35.1) | 78.5 (69.9-83.5) |
| **24** | 11.9 (8.5-14.4) | | 18.7 (14.8-21.5) | | 27.9 (21.6-32.4) | |
| **41** | 7.5 (4.9-10.0) | | 13.7 (10.7-16.6) | | 19.7 (14.8-24.4) | |
| **31*** | 13.9 (10.6-16.1) | 47.2 (37.2-53.5) | 20.9 (17.3-23.2) | 67.0 (59.3-59.3) | 31.4 (25.5-35.2) | 78.6 (70.0-83.6) |
| **39** | 8.9 (5.9-11.6) | | 15.3 (11.8-18.4) | | 22.4 (16.7-27.3) | |
| **38** | 10.5 (7.2-13.2) | | 17.1 (13.3-20.1) | | 25.3 (19.1-30.1) | |
| **34** | 13.1 (9.7-15.4) | | 20.0 (16.2-22.5) | | 30.0 (23.9-34.1) | |
| **49** | 9.3 (6.1-12.0) | | 15.8 (12.2-18.8) | | 23.1 (17.2-28.0) | |
| **25*** | 14.5 (11.4-16.5) | 49.0 (39.4-54.9) | 21.5 (18.1-23.7) | 68.4 (61.1-61.1) | 32.5 (26.8-36.0) | 80.0 (72.0-84.7) |
| **33*** | 14.0 (10.8-16.2) | 47.5 (37.7-53.8) | 21.0 (17.4-23.3) | 67.3 (59.6-59.6) | 31.6 (25.8-35.4) | 78.9 (70.5-83.9) |
| **9** | 9.5 (6.3-12.2) | | 16.0 (12.3-19.0) | | 23.4 (17.5-28.3) | |
| **27** | 12.6 (9.1-15.0) | | 19.4 (15.6-22.0) | | 29.0 (22.8-33.3) | |
| **26** | 9.2 (6.1-11.9) | | 15.6 (12.1-18.7) | | 22.9 (17.1-27.8) | |
| **8*** | 13.6 (10.3-15.8) | 46.3 (36.3-52.9) | 20.6 (16.9-23.0) | 66.4 (58.5-58.5) | 30.9 (25.0-34.8) | 77.9 (69.1-83.1) |
| **10*** | 14.9 (11.8-16.8) | 50.1 (40.8-55.8) | 21.9 (18.6-24.1) | 69.2 (62.2-62.2) | 33.1 (27.7-36.5) | 80.9 (73.3-85.4) |
| **23** | 12.8 (9.4-15.2) | | 19.7 (15.9-22.3) | | 29.4 (23.3-33.6) | |
| **22** | 10.1 (6.8-12.8) | | 16.6 (12.9-19.6) | | 24.5 (18.4-29.3) | |
| **6** | 11.0 (7.6-13.7) | | 17.7 (13.8-20.6) | | 26.3 (20.0-31.0) | |
| **20** | 9.6 (6.4-12.3) | | 16.1 (12.4-19.1) | | 23.6 (17.6-28.5) | |
| **18** | 13.2 (9.9-15.5) | | 20.1 (16.4-22.6) | | 30.2 (24.1-34.2) | |
| **13** | 10.5 (7.1-13.2) | | 17.1 (13.3-20.1) | | 25.3 (19.1-30.1) | |

| | | | | | | |
|---|---|---|---|---|---|---|
| **5** | 12.6 (9.1-15.0) | | 19.4 (15.6-22.0) | | 29.0 (22.8-33.3) | |
| **14** | 12.6 (9.2-15.0) | | 19.4 (15.6-22.1) | | 29.0 (22.9-33.3) | |
| **4** | 12.4 (9.0-14.9) | | 19.2 (15.4-21.9) | | 28.8 (22.5-33.1) | |
| **11** | 12.6 (9.2-15.0) | | 19.5 (15.6-22.1) | | 29.1 (22.9-33.4) | |
| **21*** | 13.9 (10.7-16.1) | 47.2 (37.3-53.6) | 20.9 (17.3-23.2) | 67.1 (59.3-59.3) | 31.4 (25.6-35.2) | 78.6 (70.1-83.6) |
| **19** | 10.6 (7.2-13.2) | | 17.2 (13.4-20.2) | | 25.5 (19.2-30.2) | |
| **16** | 11.3 (7.8-13.9) | | 18.0 (14.1-20.8) | | 26.7 (20.4-31.3) | |
| **2*** | 13.5 (10.2-15.8) | 46.0 (35.9-52.7) | 20.5 (16.8-22.9) | 66.2 (58.2-58.2) | 30.8 (24.8-34.7) | 77.6 (68.8-82.9) |
| **1*** | 14.8 (11.8-16.8) | 50.0 (40.7-55.7) | 21.9 (18.5-24.0) | 69.2 (62.1-62.1) | 33.1 (27.6-36.5) | 80.9 (73.2-85.3) |
| **15*** | 16.1 (13.5-17.7) | 53.9 (46.0-58.6) | 23.3 (20.4-25.1) | 72.1 (66.2-66.2) | 35.4 (30.7-38.2) | 84.1 (77.7-87.6) |
| **17** | 12.9 (9.5-15.2) | | 19.7 (15.9-22.3) | | 29.6 (23.4-33.7) | |
| **3** | 10.4 (7.0-13.0) | | 16.9 (13.2-19.9) | | 25.0 (18.9-29.9) | |
| **DSCC** | **12.7 (9.5-15.0)** | **19.0 (15.5-21.1)** | **19.5 (15.9-22.0)** | **26.2 (23.5-23.5)** | **29.2 (23.4-33.3)** | **30.7 (27.7-32.3)** |

All numbers are expressed as percentage (%) reduction relative to no intervention. Ranges within parentheses represent results from the low and high transmission models.
* Wards with high TB incidence were included in targeted interventions.

TABLE S2: **Projected percentage reduction in TB incidence in DNCC after 10 years of TB interventions.**

| *Ward* | *ACF-citywide* | *ACF-targeted* | *PT-citywide* | *PT-targeted* | *PT-targeted* | *PT-targeted* |
|---|---|---|---|---|---|---|
| **29** | 7.5 (4.9-10.1) | | 13.6 (10.6-16.6) | | 19.7 (14.8-24.5) | |
| **34*** | 11.1 (7.7-13.6) | 39.4 (28.4-47.1) | 17.7 (13.8-20.5) | 61.0 (51.8-51.8) | 26.3 (20.0-30.8) | 71.8 (61.4-78.3) |
| **35** | 7.7 (5.0-10.3) | | 13.8 (10.7-16.8) | | 20.0 (14.9-24.8) | |
| **23** | 6.1 (4.1-8.3) | | 12.0 (9.7-14.4) | | 17.0 (13.2-20.9) | |
| **36** | 6.6 (4.4-8.9) | | 12.5 (10.0-15.2) | | 17.8 (13.6-22.1) | |
| **31*** | 10.2 (6.9-12.8) | 36.3 (25.9-44.4) | 16.6 (12.9-19.5) | 58.5 (49.6-49.6) | 24.6 (18.6-29.3) | 69.0 (58.7-76.0) |
| **27** | 7.6 (5.0-10.2) | | 13.7 (10.7-16.7) | | 19.8 (14.8-24.6) | |
| **26** | 6.7 (4.4-9.0) | | 12.6 (10.0-15.3) | | 18.0 (13.7-22.3) | |
| **22** | 8.1 (5.3-10.7) | | 14.2 (11.0-17.2) | | 20.7 (15.4-25.5) | |
| **24** | 8.8 (5.8-11.5) | | 15.1 (11.6-18.1) | | 22.0 (16.4-26.9) | |
| **32** | 7.9 (5.1-10.5) | | 14.0 (10.9-16.9) | | 20.3 (15.1-25.0) | |
| **25*** | 9.8 (6.6-12.5) | 35.3 (24.8-43.6) | 16.2 (12.6-19.2) | 57.6 (48.6-48.6) | 24.0 (17.9-28.8) | 68.0 (57.5-75.3) |
| **28** | 7.2 (4.7-9.7) | | 13.2 (10.4-16.1) | | 19.0 (14.3-23.6) | |
| **20*** | 14.4 (11.3-16.3) | 49.5 (40.1-55.3) | 21.3 (17.9-23.4) | 68.8 (61.6-61.6) | 32.2 (26.7-35.6) | 80.5 (72.6-85.0) |
| **21** | 9.2 (6.1-11.8) | | 15.5 (11.9-18.5) | | 22.8 (16.9-27.6) | |
| **11** | 7.2 (4.7-9.7) | | 13.2 (10.4-16.1) | | 19.0 (14.3-23.7) | |
| **12** | 7.1 (4.7-9.6) | | 13.2 (10.3-16.0) | | 18.9 (14.2-23.5) | |
| **19** | 8.8 (5.8-11.5) | | 15.0 (11.6-18.1) | | 22.0 (16.4-26.9) | |
| **16** | 7.0 (4.6-9.4) | | 13.0 (10.2-15.7) | | 18.6 (14.1-23.1) | |
| **3** | 6.4 (4.3-8.6) | | 12.3 (9.9-14.9) | | 17.5 (13.4-21.6) | |
| **13** | 6.6 (4.4-9.0) | | 12.6 (10.0-15.2) | | 18.0 (13.7-22.2) | |
| **14** | 7.0 (4.6-9.4) | | 13.0 (10.3-15.8) | | 18.7 (14.1-23.1) | |
| **7** | 6.7 (4.4-9.0) | | 12.6 (10.0-15.3) | | 18.0 (13.7-22.3) | |
| **18*** | 9.9 (6.6-12.5) | 35.4 (24.9-43.5) | 16.3 (12.6-19.2) | 57.7 (48.7-48.7) | 24.0 (18.0-28.8) | 68.1 (57.7-75.3) |
| **4** | 7.8 (5.1-10.3) | | 13.9 (10.8-16.8) | | 20.1 (15.0-24.8) | |
| **5** | 7.8 (5.1-10.4) | | 14.0 (10.8-16.9) | | 20.2 (15.1-25.0) | |
| **17** | 8.0 (5.2-10.6) | | 14.1 (11.0-17.1) | | 20.5 (15.3-25.3) | |
| **2** | 8.3 (5.4-10.9) | | 14.5 (11.2-17.5) | | 21.1 (15.7-26.0) | |
| **15*** | 9.6 (6.4-12.2) | 34.4 (24.0-42.7) | 15.9 (12.3-18.9) | 56.9 (47.9-47.9) | 23.5 (17.5-28.3) | 67.2 (56.7-74.5) |
| **6** | 7.3 (4.8-9.8) | | 13.3 (10.5-16.2) | | 19.2 (14.4-23.8) | |
| **1** | 8.3 (5.4-10.9) | | 14.5 (11.2-17.4) | | 21.1 (15.7-25.9) | |
| **33** | 7.2 (4.7-9.7) | | 13.3 (10.4-16.1) | | 19.1 (14.4-23.7) | |
| **30** | 6.9 (4.5-9.3) | | 12.9 (10.2-15.6) | | 18.4 (14.0-22.8) | |
| **10** | 8.4 (5.5-11.0) | | 14.6 (11.3-17.5) | | 21.2 (15.8-26.1) | |
| **9** | 6.8 (4.5-9.2) | | 12.8 (10.1-15.5) | | 18.3 (13.9-22.7) | |
| **8** | 9.2 (6.1-11.9) | | 15.5 (12.0-18.5) | | 22.8 (17.0-27.7) | |
| **DNCC** | **9.5 (6.6-12.0)** | **17.3 (13.0-20.3)** | **15.8 (12.5-18.6)** | **26.1 (22.6-22.6)** | **23.3 (17.8-27.8)** | **30.7 (26.7-33.2)** |

All numbers are expressed as percentage (%) reduction relative to no intervention. Ranges within parentheses represent results from the low and high transmission models.
* Wards with high TB incidence were included in targeted interventions.

TABLE S3: **Projected percentage reduction in TB incidence in DSCC after 5 years of TB interventions.**

| *Ward* | *ACF--citywide* | *ACF--targeted* | *PT--citywide* | *PT--targeted* | *Combination-citywide* | *Combination--targeted* |
|---|---|---|---|---|---|---|
| **44** | 5.7 (3.9-7.3) | | 11.5 (9.5-13.3) | | 16.3 (12.8-19.4) | |
| **54** | 5.1 (3.5-6.7) | | 10.9 (9.1-12.6) | | 15.3 (12.1-18.2) | |
| **53** | 6.7 (4.7-8.3) | | 12.7 (10.4-14.5) | | 18.4 (14.4-21.4) | |
| **52** | 5.2 (3.5-6.8) | | 11.0 (9.1-12.8) | | 15.4 (12.2-18.4) | |
| **47** | 5.1 (3.4-6.5) | | 10.8 (9-12.5) | | 15.1 (12.0-18.0) | |
| **46** | 5.2 (3.5-6.7) | | 10.9 (9.1-12.6) | | 15.3 (12.1-18.2) | |
| **12** | 8.1 (6.0-9.5) | | 14.3 (11.9-15.8) | | 21.0 (17.0-23.6) | |
| **43** | 4.9 (3.3-6.3) | | 10.6 (8.9-12.3) | | 14.8 (11.8-17.6) | |
| **45** | 6.8 (4.7-8.4) | | 12.8 (10.4-14.5) | | 18.5 (14.4-21.5) | |
| **51** | 6.1 (4.1-7.7) | | 12.0 (9.8-13.8) | | 17.1 (13.3-20.1) | |
| **37** | 5.0 (3.4-6.5) | | 10.8 (9.0-12.4) | | 15.0 (11.9-17.8) | |
| **50*** | 10.2 (8.7-11.0) | 39.0 (33.6-42.0) | 16.5 (14.9-17.4) | 59.2 (54.9-61.4) | 24.9 (22.1-26.4) | 73.7 (68.0-76.7) |
| **42** | 6.9 (4.7-8.4) | | 12.9 (10.5-14.6) | | 18.6 (14.5-21.6) | |
| **7** | 7.8 (5.7-9.3) | | 13.9 (11.6-15.5) | | 20.4 (16.4-23.2) | |
| **36** | 7.5 (5.4-9.0) | | 13.6 (11.2-15.3) | | 19.9 (15.8-22.7) | |
| **30** | 8.9 (6.9-10.1) | | 15.1 (12.9-16.5) | | 22.5 (18.8-24.8) | |
| **32** | 8.3 (6.2-9.7) | | 14.5 (12.2-16.0) | | 21.4 (17.5-24.0) | |
| **48** | 8.1 (6.0-9.5) | | 14.2 (11.9-15.8) | | 21.0 (17.0-23.6) | |
| **40** | 9.1 (7.2-10.3) | | 15.4 (13.3-16.6) | | 22.9 (19.3-25.1) | |
| **29** | 8.9 (7.0-10.1) | | 15.2 (13.0-16.5) | | 22.6 (18.9-24.8) | |
| **35*** | 10.8 (9.8-11.3) | 41.8 (38.1-43.7) | 17.3 (16.2-17.8) | 61.3 (58.5-62.8) | 26.2 (24.4-27.2) | 76.8 (73.1-78.8) |
| **28*** | 9.6 (7.9-10.6) | 36.9 (30.7-40.6) | 15.9 (14.0-17.0) | 57.5 (52.5-60.3) | 23.9 (20.6-25.8) | 71.4 (64.8-75.2) |
| **24** | 8.7 (6.7-9.9) | | 14.9 (12.6-16.3) | | 22.1 (18.2-24.4) | |
| **41** | 5.8 (3.9-7.4) | | 11.7 (9.6-13.5) | | 16.6 (12.9-19.6) | |
| **31*** | 9.6 (7.9-10.6) | 37.0 (30.8-40.7) | 15.9 (14.1-17.0) | 57.5 (52.6-60.4) | 23.9 (20.7-25.8) | 71.5 (64.9-75.2) |
| **39** | 6.8 (4.7-8.4) | | 12.9 (10.5-14.6) | | 18.6 (14.5-21.6) | |
| **38** | 7.9 (5.7-9.3) | | 14.0 (11.6-15.6) | | 20.5 (16.5-23.3) | |
| **34** | 9.2 (7.4-10.4) | | 15.5 (13.5-16.7) | | 23.2 (19.7-25.3) | |
| **49** | 7.1 (5.0-8.7) | | 13.1 (10.7-14.9) | | 19.1 (15.0-22.0) | |
| **25*** | 9.9 (8.3-10.8) | 38.0 (32.2-41.4) | 16.2 (14.5-17.2) | 58.3 (53.8-60.9) | 24.4 (21.4-26.1) | 72.6 (66.5-76.0) |
| **33*** | 9.7 (8.0-10.7) | 37.2 (31.1-40.8) | 16.0 (14.1-17.1) | 57.7 (52.8-60.5) | 24.0 (20.8-25.9) | 71.7 (65.2-75.4) |
| **9** | 7.2 (5.1-8.8) | | 13.3 (10.9-15.0) | | 19.3 (15.2-22.2) | |
| **27** | 9.0 (7.1-10.2) | | 15.2 (13.1-16.5) | | 22.7 (19.0-24.9) | |
| **26** | 7.0 (4.9-8.6) | | 13.1 (10.7-14.8) | | 19.0 (14.9-21.9) | |
| **8*** | 9.5 (7.7-10.5) | 36.5 (30.1-40.3) | 15.8 (13.9-16.9) | 57.2 (52.0-60.1) | 23.7 (20.3-25.6) | 71.0 (64.1-74.9) |
| **10*** | 10.0 (8.5-10.9) | 38.6 (33.1-41.8) | 16.4 (14.7-17.4) | 58.8 (54.5-61.2) | 24.7 (21.9-26.3) | 73.2 (67.4-76.4) |
| **23** | 9.1 (7.2-10.2) | | 15.4 (13.3-16.6) | | 22.9 (19.3-25.1) | |
| **22** | 7.6 (5.5-9.1) | | 13.7 (11.3-15.3) | | 20.0 (15.9-22.8) | |
| **6** | 8.2 (6.1-9.5) | | 14.3 (12.0-15.9) | | 21.1 (17.1-23.7) | |
| **20** | 7.3 (5.1-8.8) | | 13.3 (10.9-15.0) | | 19.4 (15.3-22.3) | |
| **18** | 9.3 (7.5-10.4) | | 15.6 (13.6-16.8) | | 23.3 (19.8-25.4) | |
| **13** | 7.8 (5.7-9.3) | | 14.0 (11.6-15.6) | | 20.5 (16.4-23.2) | |
| **5** | 9.0 (7.1-10.2) | | 15.2 (13.1-16.5) | | 22.7 (19.0-24.9) | |
| **14** | 9.0 (7.1-10.2) | | 15.2 (13.1-16.5) | | 22.7 (19.1-24.9) | |
| **4** | 8.9 (7.0-10.1) | | 15.1 (13.0-16.5) | | 22.5 (18.8-24.8) | |
| **11** | 9.0 (7.1-10.2) | | 15.3 (13.1-16.6) | | 22.7 (19.1-24.9) | |
| **21*** | 9.6 (7.9-10.6) | 37.0 (30.8-40.7) | 16.0 (14.1-17.1) | 57.6 (52.6-60.4) | 23.9 (20.7-25.8) | 71.5 (64.9-75.2) |
| **19** | 7.9 (5.8-9.3) | | 14.0 (11.7-15.6) | | 20.6 (16.6-23.3) | |
| **16** | 8.3 (6.2-9.6) | | 14.5 (12.1-16.0) | | 21.4 (17.4-23.9) | |

| | | | | | | |
|---|---|---|---|---|---|---|
| **2*** | 9.4 (7.7-10.5) | 36.4 (29.9-40.2) | 15.8 (13.8-16.9) | 57.0 (51.8-60.0) | 23.6 (20.2-25.6) | 70.8 (63.9-74.7) |
| **1*** | 10.0 (8.5-10.9) | 38.6 (33.0-41.7) | 16.4 (14.7-17.4) | 58.8 (54.4-61.2) | 24.7 (21.8-26.3) | 73.2 (67.4-76.4) |
| **15*** | 10.5 (9.3-11.2) | 40.6 (36.1-43.1) | 17.0 (15.6-17.7) | 60.4 (56.9-62.2) | 25.7 (23.4-26.9) | 75.4 (70.8-77.9) |
| **17** | 9.1 (7.3-10.3) | | 15.4 (13.3-16.7) | | 23.0 (19.4-25.1) | |
| **3** | 7.8 (5.6-9.2) | | 13.9 (11.5-15.5) | | 20.4 (16.3-23.1) | |
| **DSCC** | **8.9 (7.1-10.1)** | **14.6 (12.5-15.8)** | **15.2 (13.2-16.5)** | **22.3 (20.6-23.2)** | **22.6 (19.2-24.8)** | **27.7 (25.5-28.9)** |

All numbers are expressed as percentage (%) reduction relative to no intervention. Ranges within parentheses represent results from the low and high transmission models.
* Wards with high TB incidence were included in targeted interventions.

TABLE S4: **Projected percentage reduction in TB incidence in DNCC after 5 years of TB interventions.**

| *Ward* | *ACF-citywide* | *ACF-targeted* | *PT-citywide* | *PT-targeted* | *Combination-citywide* | *Combination-targeted* |
|---|---|---|---|---|---|---|
| **29** | 5.8 (4.0-7.4) | | 11.6 (9.5-13.4) | | 16.6 (12.9-19.6) | |
| **34*** | 8.1 (6.1-9.5) | 32.1 (24.4-37) | 14.2 (11.9-15.7) | 53.7 (47.2-57.5) | 21.0 (17.1-23.5) | 66.2 (57.6-71.3) |
| **35** | 6.0 (4.0-7.6) | | 11.7 (9.6-13.5) | | 16.8 (13.1-19.8) | |
| **23** | 4.7 (3.2-6.1) | | 10.3 (8.6-11.9) | | 14.3 (11.4-17.0) | |
| **36** | 5.1 (3.4-6.6) | | 10.7 (8.9-12.4) | | 15.1 (11.9-17.9) | |
| **31*** | 7.6 (5.5-9.0) | 30.0 (22.2-35.3) | 13.6 (11.3-15.2) | 51.9 (45.4-56.2) | 20.0 (16.0-22.7) | 63.9 (55.1-69.5) |
| **27** | 5.9 (4.0-7.5) | | 11.7 (9.5-13.5) | | 16.6 (13.0-19.7) | |
| **26** | 5.1 (3.5-6.7) | | 10.8 (8.9-12.5) | | 15.2 (12.0-18.1) | |
| **22** | 6.2 (4.3-7.8) | | 12.0 (9.8-13.8) | | 17.3 (13.5-20.4) | |
| **24** | 6.7 (4.7-8.3) | | 12.6 (10.3-14.4) | | 18.3 (14.3-21.3) | |
| **32** | 6.1 (4.1-7.7) | | 11.9 (9.7-13.7) | | 17.0 (13.2-20.0) | |
| **25*** | 7.4 (5.3-8.9) | 29.3 (21.3-34.8) | 13.4 (11.0-15.0) | 51.3 (44.6-55.8) | 19.6 (15.5-22.4) | 63.0 (53.9-68.9) |
| **28** | 5.6 (3.8-7.1) | | 11.3 (9.3-13.1) | | 16.0 (12.5-19.1) | |
| **20*** | 9.8 (8.2-10.6) | 38.3 (32.6-41.6) | 16.0 (14.3-16.9) | 58.5 (54.1-61.0) | 24.1 (21.2-25.7) | 72.9 (66.9-76.2) |
| **21** | 7.0 (4.9-8.5) | | 12.9 (10.5-14.6) | | 18.8 (14.7-21.7) | |
| **11** | 5.6 (3.8-7.2) | | 11.3 (9.2-13.1) | | 16.0 (12.5-19.1) | |
| **12** | 5.5 (3.7-7.1) | | 11.3 (9.2-13.0) | | 15.9 (12.4-19.0) | |
| **19** | 6.7 (4.7-8.3) | | 12.6 (10.3-14.4) | | 18.3 (14.3-21.3) | |
| **16** | 5.4 (3.7-6.9) | | 11.1 (9.1-12.8) | | 15.7 (12.3-18.7) | |
| **3** | 4.9 (3.4-6.4) | | 10.6 (8.8-12.2) | | 14.8 (11.7-17.6) | |
| **13** | 5.1 (3.5-6.6) | | 10.8 (8.9-12.5) | | 15.2 (11.9-18.1) | |
| **14** | 5.4 (3.7-7.0) | | 11.1 (9.1-12.9) | | 15.7 (12.3-18.7) | |
| **7** | 5.2 (3.5-6.7) | | 10.8 (9.0-12.5) | | 15.2 (12.0-18.1) | |
| **18*** | 7.4 (5.3-8.9) | 29.3 (21.5-34.8) | 13.4 (11.1-15) | 51.4 (44.7-55.7) | 19.6 (15.6-22.4) | 63.1 (54.1-68.9) |
| **4** | 6.0 (4.1-7.6) | | 11.8 (9.6-13.6) | | 16.8 (13.1-19.9) | |
| **5** | 6.0 (4.1-7.6) | | 11.8 (9.6-13.6) | | 17.0 (13.2-20.0) | |
| **17** | 6.2 (4.2-7.7) | | 12.0 (9.7-13.7) | | 17.2 (13.4-20.2) | |
| **2** | 6.4 (4.4-8.0) | | 12.2 (10.0-14.0) | | 17.6 (13.7-20.6) | |
| **15*** | 7.2 (5.1-8.7) | 28.6 (20.7-34.2) | 13.2 (10.8-14.8) | 50.8 (44.1-55.3) | 19.3 (15.2-22.1) | 62.3 (53.2-68.3) |
| **6** | 5.6 (3.8-7.2) | | 11.4 (9.3-13.1) | | 16.2 (12.6-19.2) | |
| **1** | 6.4 (4.4-7.9) | | 12.2 (9.9-14.0) | | 17.6 (13.7-20.6) | |
| **33** | 5.6 (3.8-7.2) | | 11.3 (9.3-13.1) | | 16.1 (12.6-19.1) | |
| **30** | 5.3 (3.6-6.8) | | 11.0 (9.1-12.7) | | 15.5 (12.2-18.5) | |
| **10** | 6.4 (4.4-8.0) | | 12.3 (10.0-14.0) | | 17.7 (13.8-20.7) | |
| **9** | 5.3 (3.6-6.8) | | 11.0 (9.0-12.7) | | 15.4 (12.1-18.4) | |
| **8** | 7.0 (4.9-8.5) | | 12.9 (10.6-14.6) | | 18.8 (14.8-21.7) | |
| **DNCC** | **7.0 (5.1-8.5)** | **13.9 (10.9-15.8)** | **13.0 (10.8-14.5)** | **22.8 (20.4-24.3)** | **18.9 (15.2-21.6)** | **28.2 (24.9-30.2)** |

All numbers are expressed as percentage (%) reduction relative to no intervention. Ranges within parentheses represent results from the low and high transmission models.
* Wards with high TB incidence were included in targeted interventions.

FIGURE S1: **Impact of TB interventions on ward-level TB incidence in DSCC after 20 years of intervention in the low-transmission scenario.** The colors for each ward show the projected TB incidence after 20 years of intervention, and the bubbles indicate the absolute size of the reductions (the reduction in the number of incident TB cases achieved by the intervention in year 20). Panel (a) represents city-wide active case finding, (b) targeted case finding, (c) city-wide preventive therapy, (d) targeted preventive therapy, (e) combination of citywide active case finding and preventive therapy, and (f) combination of targeted active case finding and preventive therapy.

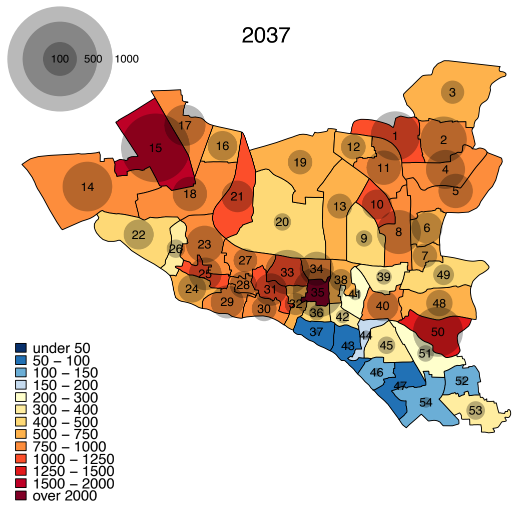

(a)

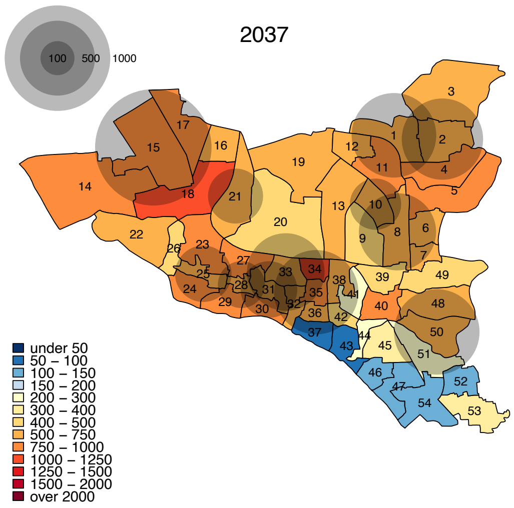

(b)

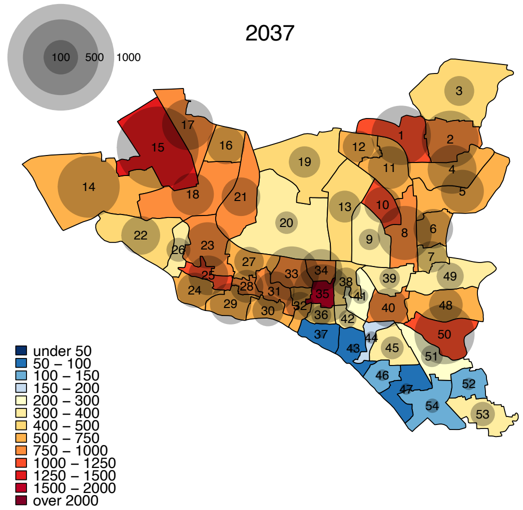

(c)

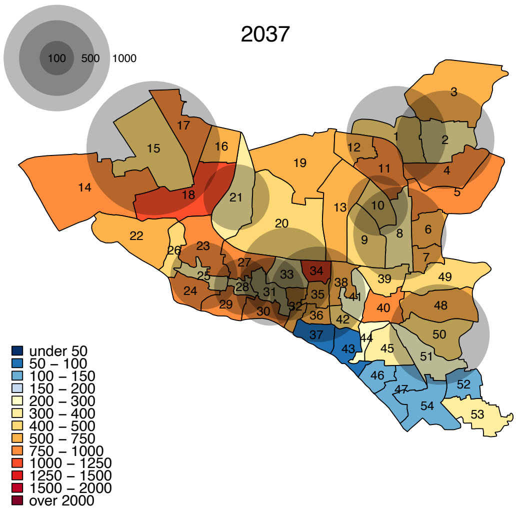

(d)

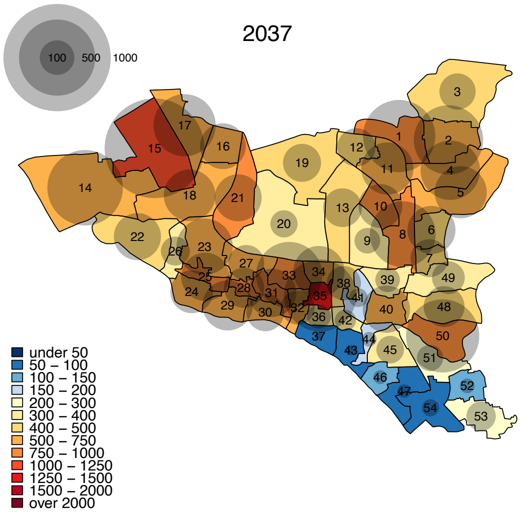

(e)

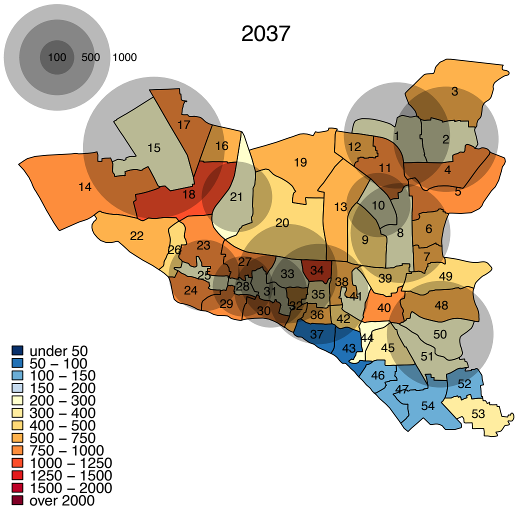

(f)

FIGURE S2: **Impact of TB interventions on ward-level TB incidence in DNCC after 20 years.** The colors for each ward show the projected TB incidence after 20 years of intervention, and the bubbles indicate the absolute size of the reductions (the reduction in the number of incident TB cases achieved by the intervention in year 20). Panel (a) represents city-wide active case finding, (b) targeted case finding, (c) city-wide preventive therapy, (d) targeted preventive therapy, (e) combination of citywide active case finding and preventative therapy, and (f) combination of targeted active case finding and preventative therapy.

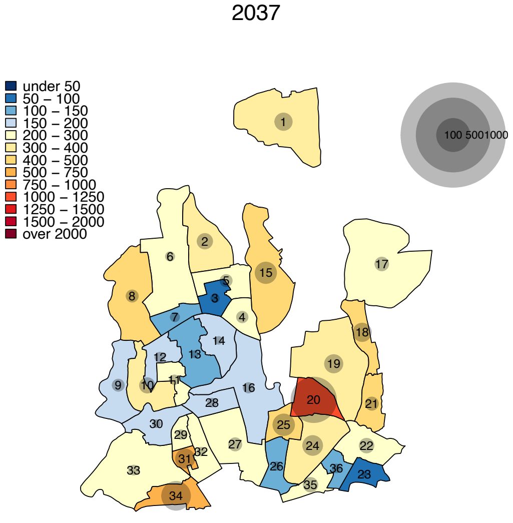

(a)

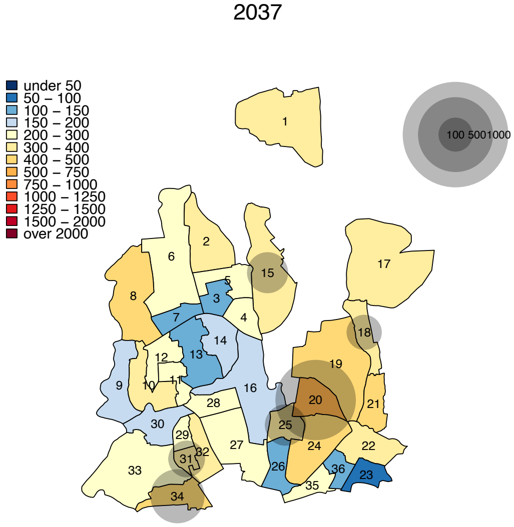

(b)

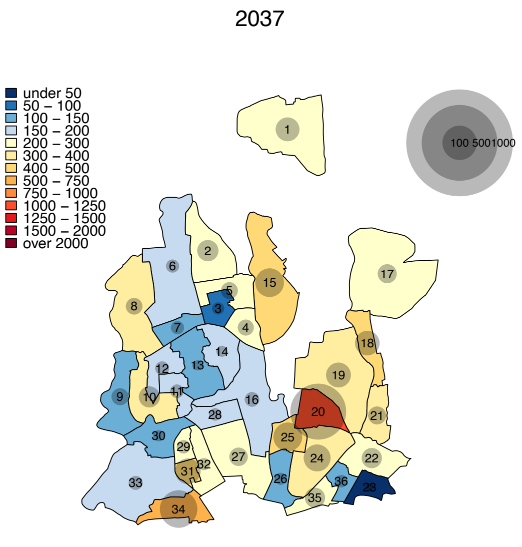
2037
under 50
50 – 100
100 – 150
150 – 200
200 – 300
300 – 400
400 – 500
500 – 750
750 – 1000
1000 – 1250
1250 – 1500
1500 – 2000
over 2000
100 5001000

(c)

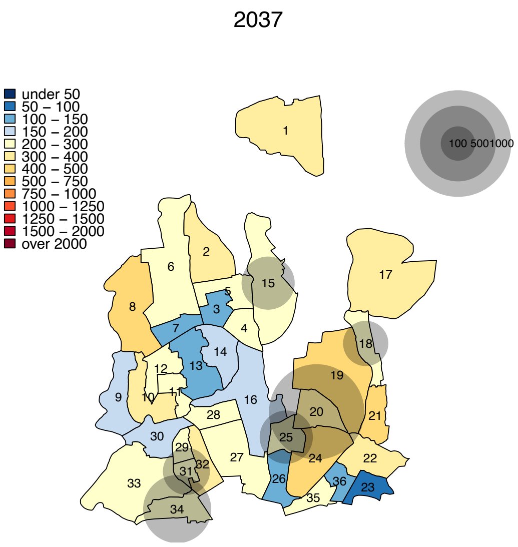
2037
under 50
50 – 100
100 – 150
150 – 200
200 – 300
300 – 400
400 – 500
500 – 750
750 – 1000
1000 – 1250
1250 – 1500
1500 – 2000
over 2000
100 5001000

(d)

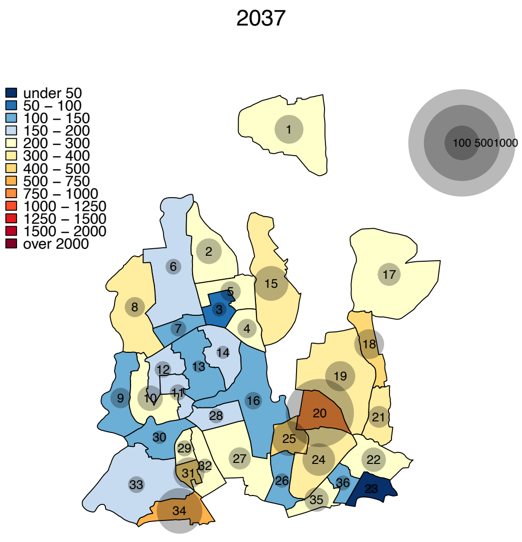
2037
under 50
50 – 100
100 – 150
150 – 200
200 – 300
300 – 400
400 – 500
500 – 750
750 – 1000
1000 – 1250
1250 – 1500
1500 – 2000
over 2000
100 5001000

(e)

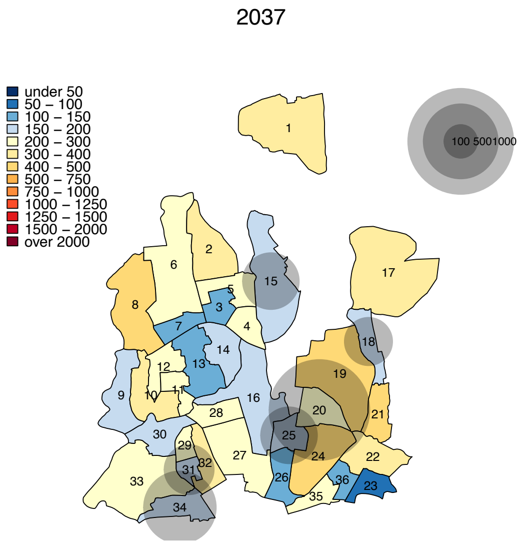
2037
under 50
50 – 100
100 – 150
150 – 200
200 – 300
300 – 400
400 – 500
500 – 750
750 – 1000
1000 – 1250
1250 – 1500
1500 – 2000
over 2000
100 5001000

(f)

FIGURE S3: **Impact of TB interventions on ward-level TB incidence in DSCC after 10 years of intervention in the low-transmission scenario.** The colors for each ward show the projected TB incidence after 10 years of intervention, and the bubbles indicate the absolute size of the reductions (the reduction in the number of incident TB cases achieved by the intervention in year 10). Panel (a) represents city-wide active case finding, (b) targeted case finding, (c) city-wide preventive therapy, (d) targeted preventive therapy, (e) combination of citywide active case finding and preventive therapy, and (f) combination of targeted active case finding and preventive therapy.

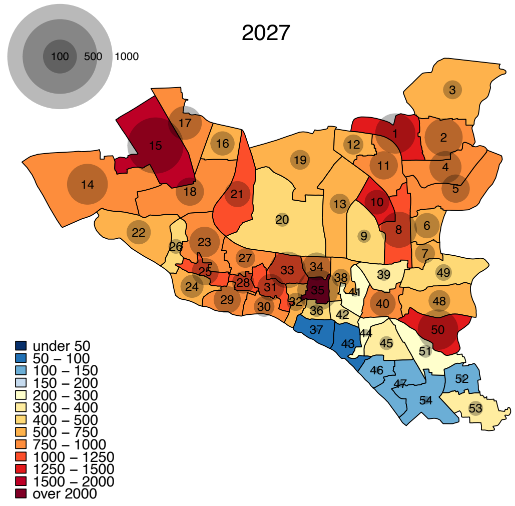

(a)

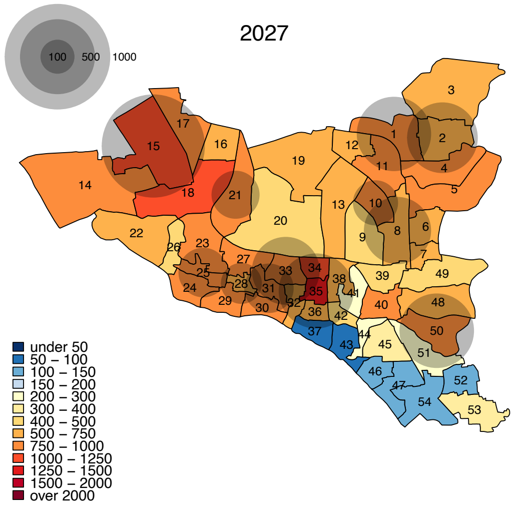

(b)

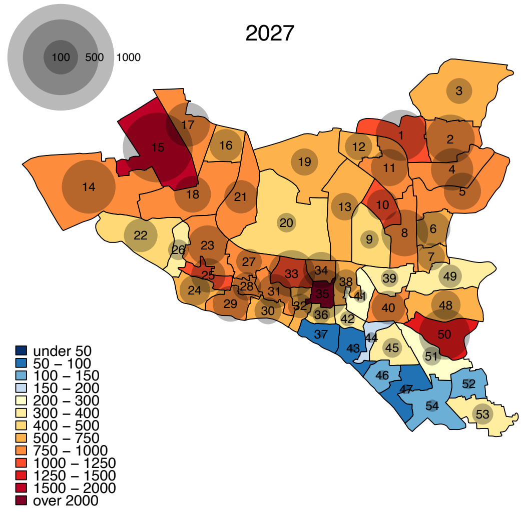

(c)

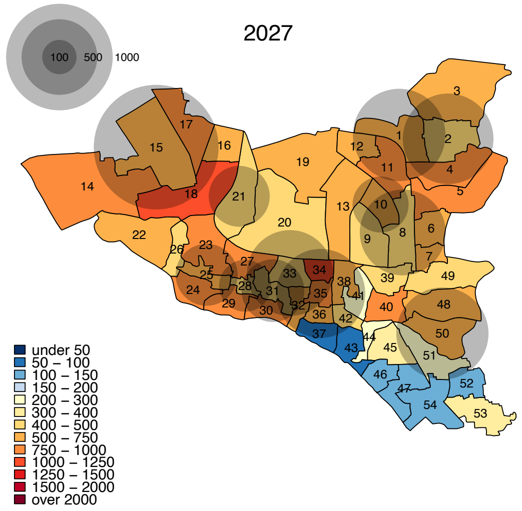

(d)

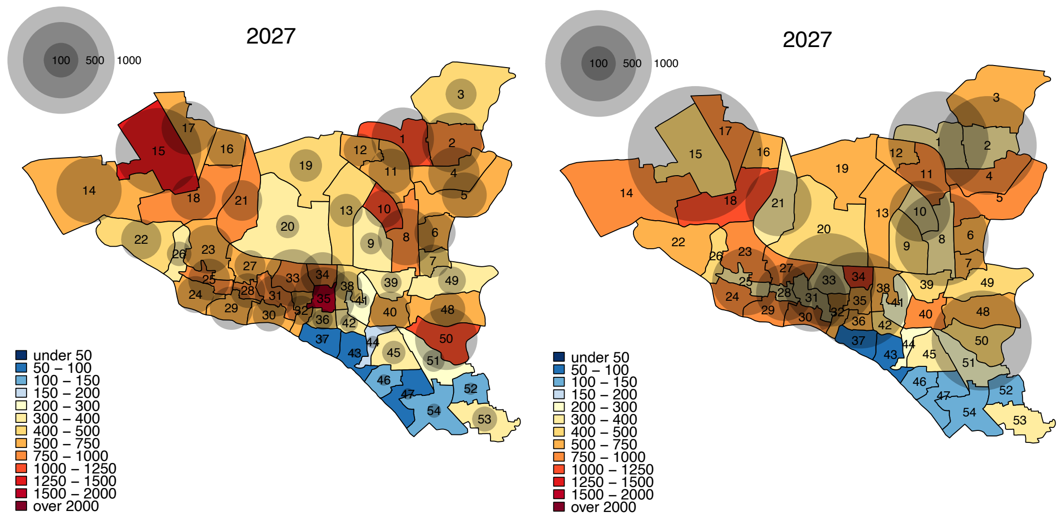

(e) (f)

FIGURE S4: **Impact of TB interventions on ward-level TB incidence in DSCC after 10 years of intervention in the high-transmission scenario.** The colors for each ward show the projected TB incidence after 10 years of intervention, and the bubbles indicate the absolute size of the reductions (the reduction in the number of incident TB cases achieved by the intervention in year 10). Panel (a) represents city-wide active case finding, (b) targeted case finding, (c) city-wide preventive therapy, (d) targeted preventive therapy, (e) combination of citywide active case finding and preventive therapy, and (f) combination of targeted active case finding and preventive therapy.

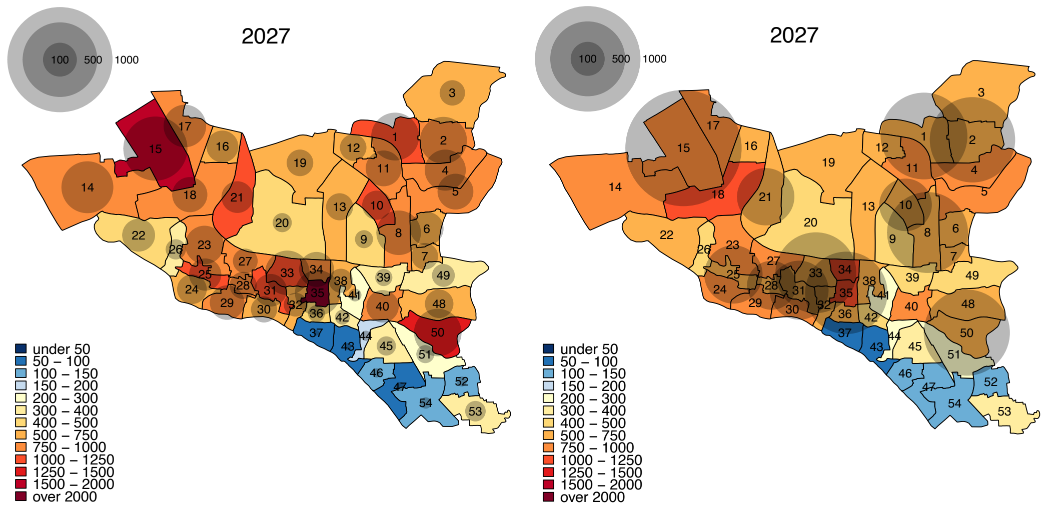

(a) (b)

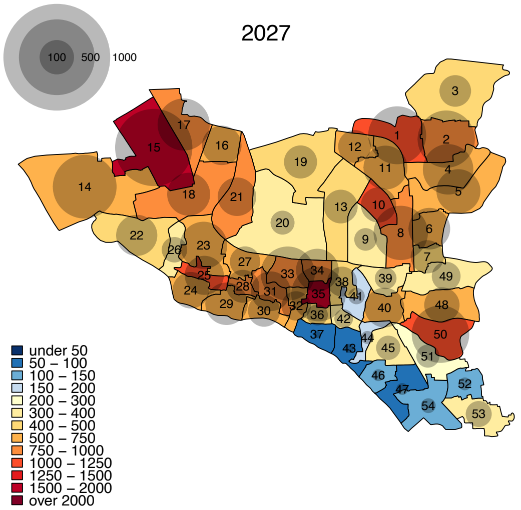
2027
100
500
1000
under 50
50 – 100
100 – 150
150 – 200
200 – 300
300 – 400
400 – 500
500 – 750
750 – 1000
1000 – 1250
1250 – 1500
1500 – 2000
over 2000

(c)

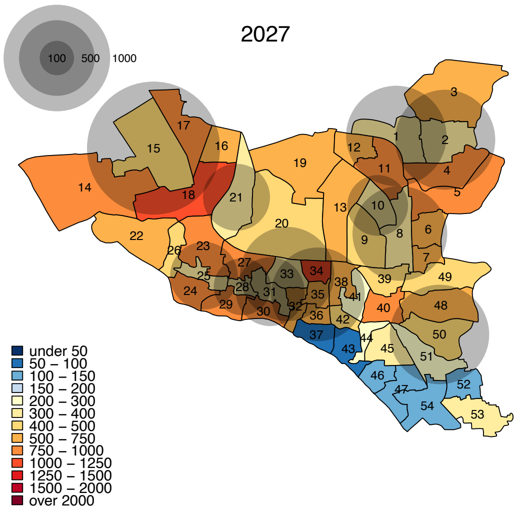
2027
100
500
1000
under 50
50 – 100
100 – 150
150 – 200
200 – 300
300 – 400
400 – 500
500 – 750
750 – 1000
1000 – 1250
1250 – 1500
1500 – 2000
over 2000

(d)

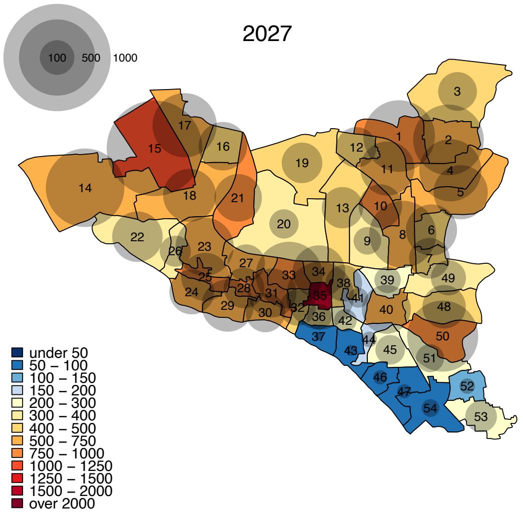
2027
100
500
1000
under 50
50 – 100
100 – 150
150 – 200
200 – 300
300 – 400
400 – 500
500 – 750
750 – 1000
1000 – 1250
1250 – 1500
1500 – 2000
over 2000

(e)

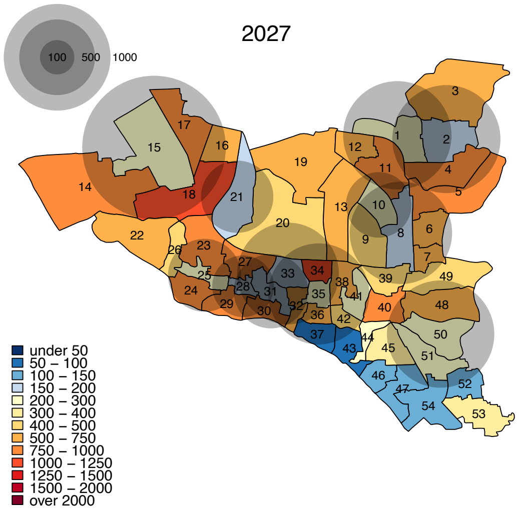
2027
100
500
1000
under 50
50 – 100
100 – 150
150 – 200
200 – 300
300 – 400
400 – 500
500 – 750
750 – 1000
1000 – 1250
1250 – 1500
1500 – 2000
over 2000

(f)

FIGURE S5: **Impact of TB interventions on ward-level TB incidence in DNCC after 10 years of intervention in the low-transmission scenario.** The colors for each ward show the projected TB incidence after 10 years of intervention, and the bubbles indicate the absolute size of the reductions (the reduction in the number of incident TB cases achieved by the intervention in year 10). Panel (a) represents city-wide active case finding, (b) targeted case finding, (c) city-wide preventive therapy, (d) targeted preventive therapy, (e) combination of citywide active case finding and preventive therapy, and (f) combination of targeted active case finding and preventive therapy.

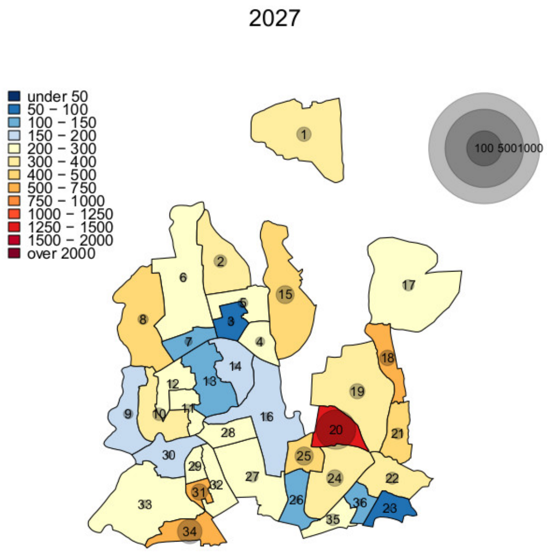

(a)

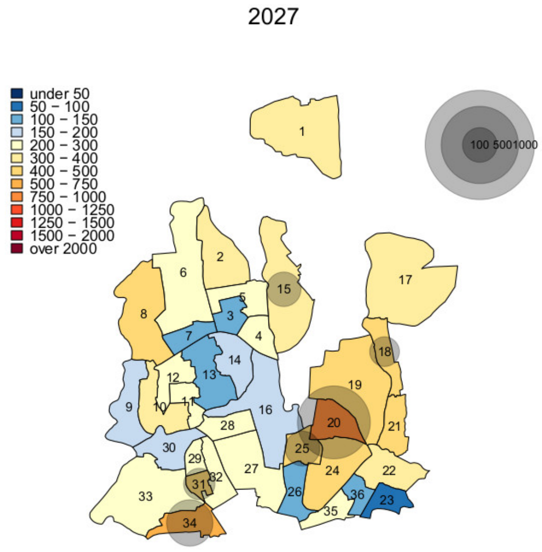

(b)

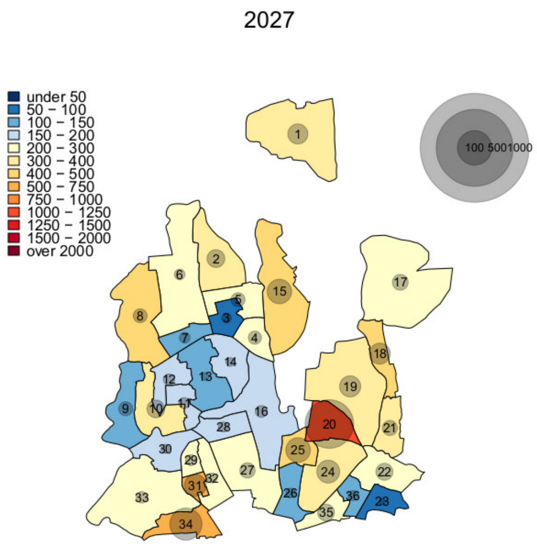

(c)

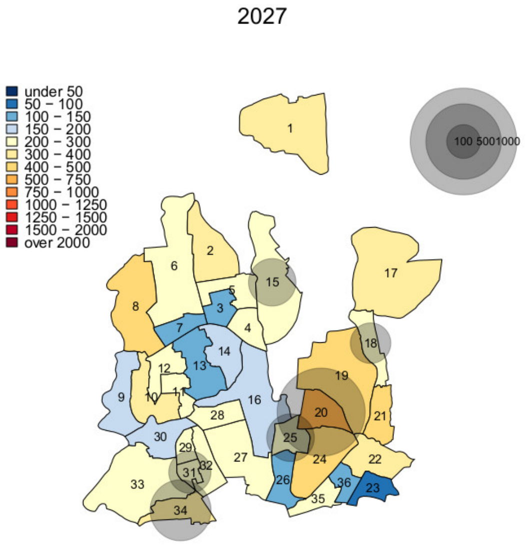

(d)

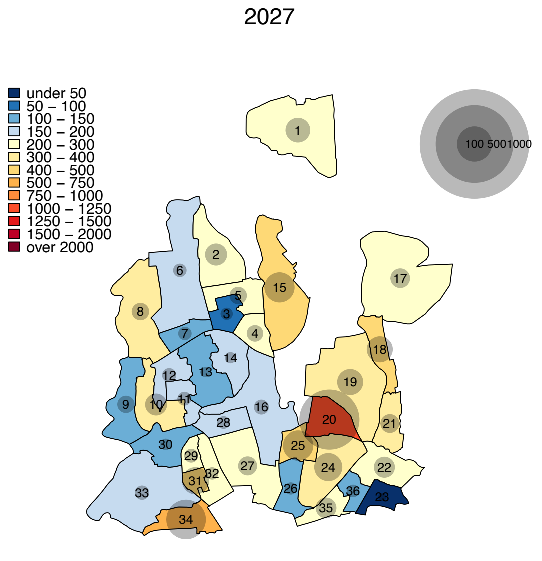
2027
under 50
50 – 100
100 – 150
150 – 200
200 – 300
300 – 400
400 – 500
500 – 750
750 – 1000
1000 – 1250
1250 – 1500
1500 – 2000
over 2000
100 5001000

(e)

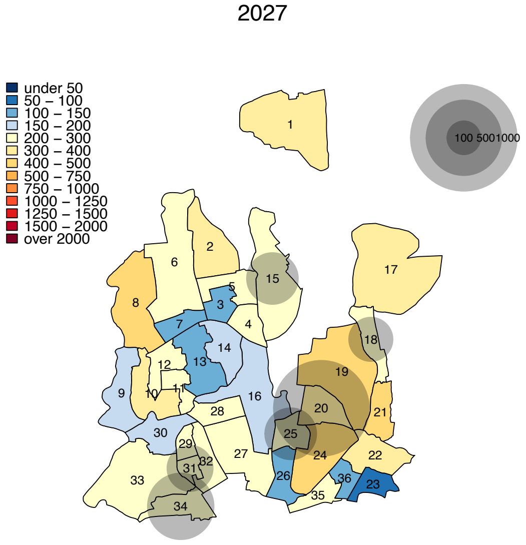
2027
under 50
50 – 100
100 – 150
150 – 200
200 – 300
300 – 400
400 – 500
500 – 750
750 – 1000
1000 – 1250
1250 – 1500
1500 – 2000
over 2000
100 5001000

(f)

FIGURE S6: **Impact of TB interventions on ward-level TB incidence in DNCC after 10 years of intervention in the high-transmission scenario.** The colors for each ward show the projected TB incidence after 10 years of intervention, and the bubbles indicate the absolute size of the reductions (the reduction in the number of incident TB cases achieved by the intervention in year 10). Panel (a) represents city-wide active case finding, (b) targeted case finding, (c) city-wide preventive therapy, (d) targeted preventive therapy, (e) combination of citywide active case finding and preventive therapy, and (f) combination of targeted active case finding and preventive therapy.

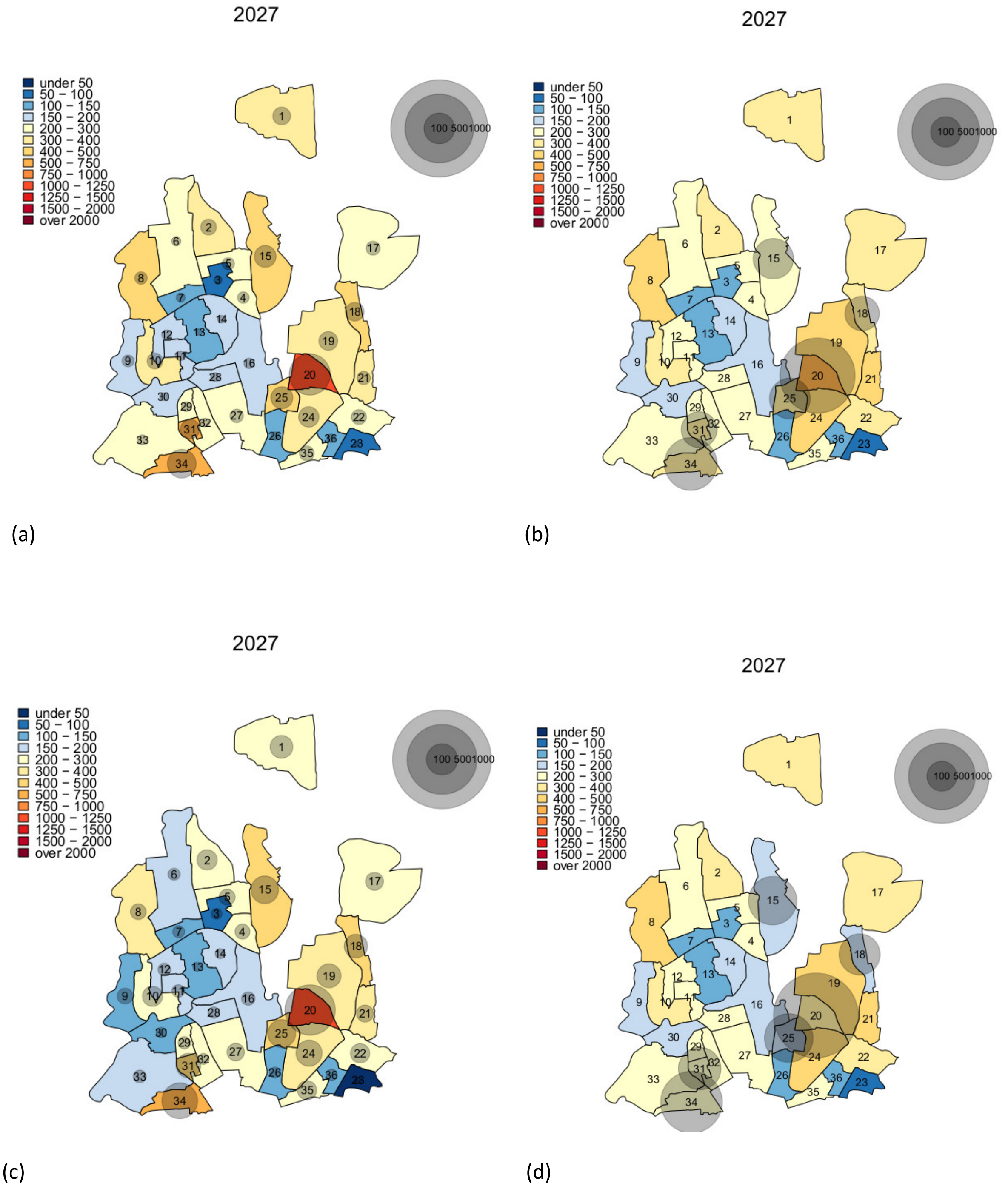

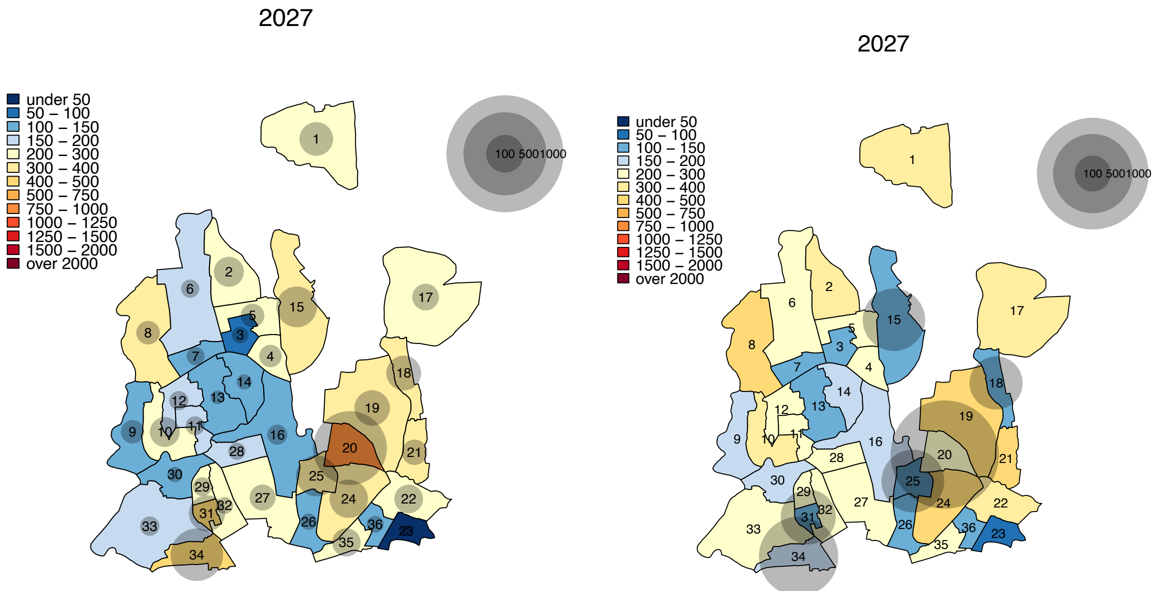

(e) (f)

FIGURE S7: **Impact of TB interventions on ward-level TB incidence in DSCC after 10 years.** The colors for each ward show the projected TB incidence after 10 years of intervention, and the bubbles indicate the absolute size of the reductions (the reduction in the number of incident TB cases achieved by the intervention in year 10). Panel (a) represents city-wide active case finding, (b) targeted case finding, (c) city-wide preventive therapy, (d) targeted preventive therapy, (e) combination of citywide active case finding and preventive therapy, and (f) combination of targeted active case finding and preventive therapy.

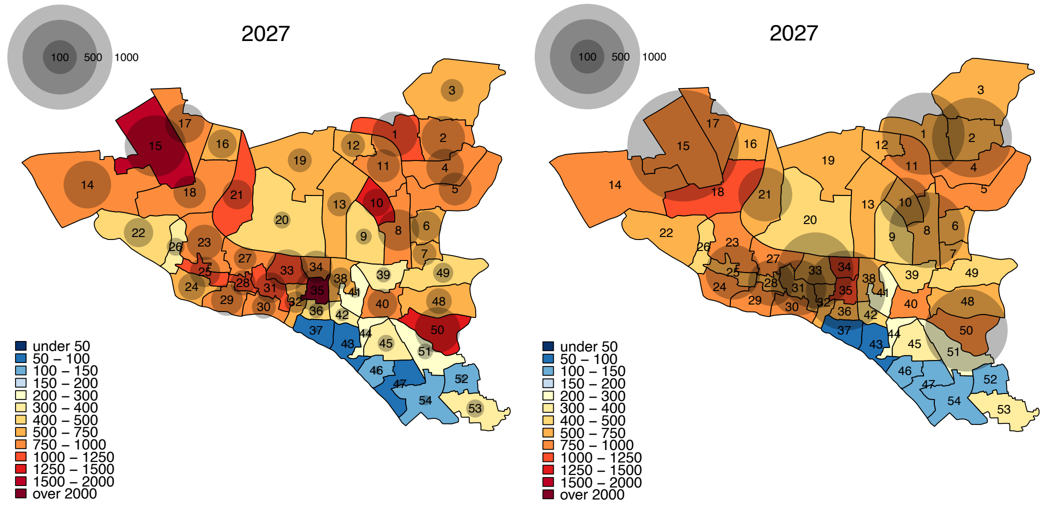

(a) (b)

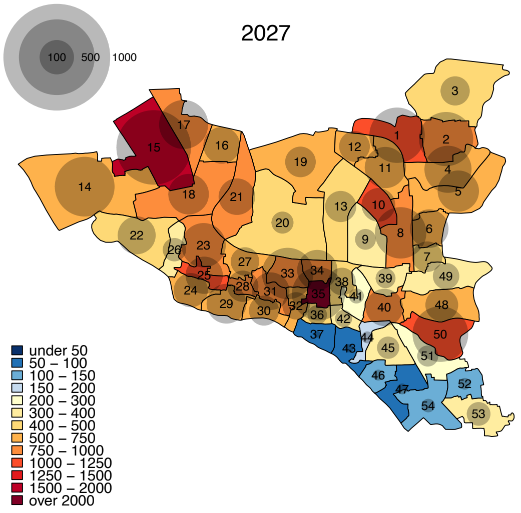

(c)

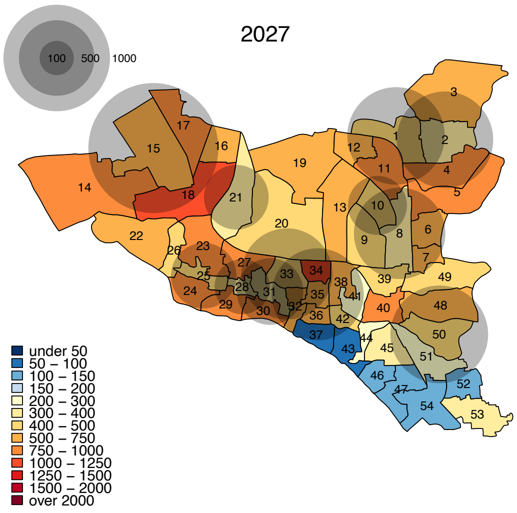

(d)

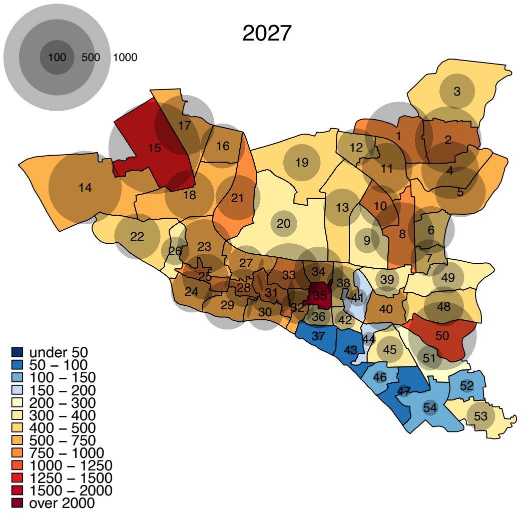

(e)

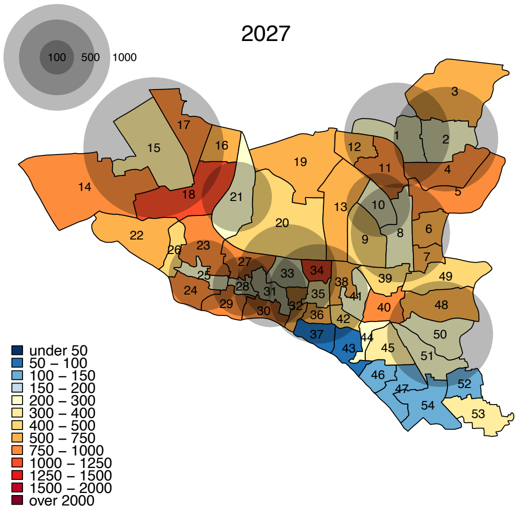

(f)

FIGURE S8: **Impact of TB interventions on ward-level TB incidence in DNCC after 10 years.** The colors for each ward show the projected TB incidence after 10 years of intervention, and the bubbles indicate the absolute size of the reductions (the reduction in the number of incident TB cases achieved by the intervention in year 10). Panel (a) represents city-wide active case finding, (b) targeted case finding, (c) city-wide preventive therapy, (d) targeted preventive therapy, (e) combination of citywide active case finding and preventive therapy, and (f) combination of targeted active case finding and preventive therapy.

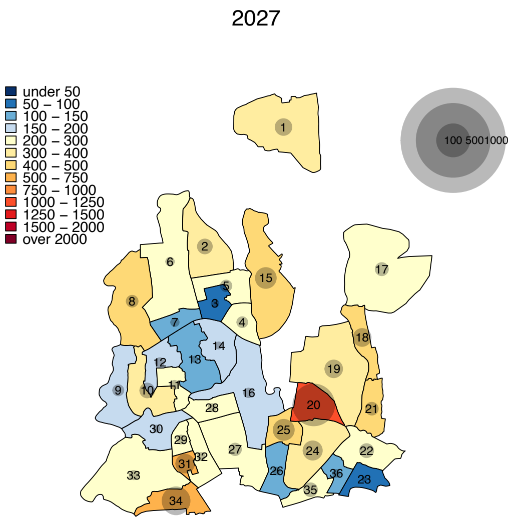
2027
under 50
50 – 100
100 – 150
150 – 200
200 – 300
300 – 400
400 – 500
500 – 750
750 – 1000
1000 – 1250
1250 – 1500
1500 – 2000
over 2000
100 5001000

(a)

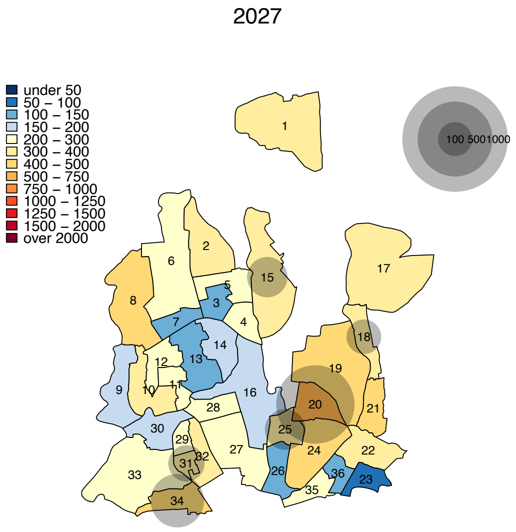
2027
under 50
50 – 100
100 – 150
150 – 200
200 – 300
300 – 400
400 – 500
500 – 750
750 – 1000
1000 – 1250
1250 – 1500
1500 – 2000
over 2000
100 5001000

(b)

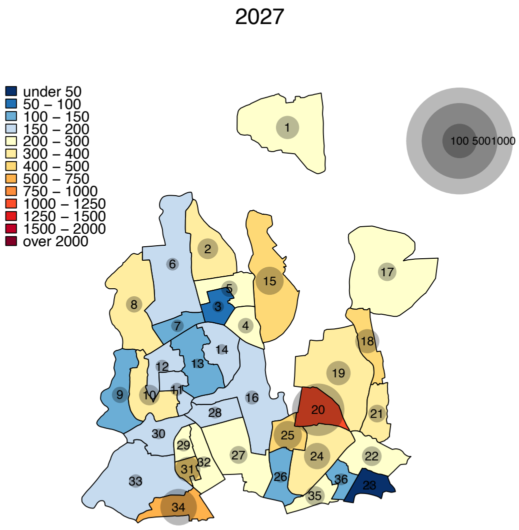
2027
under 50
50 – 100
100 – 150
150 – 200
200 – 300
300 – 400
400 – 500
500 – 750
750 – 1000
1000 – 1250
1250 – 1500
1500 – 2000
over 2000
100 5001000

(c)

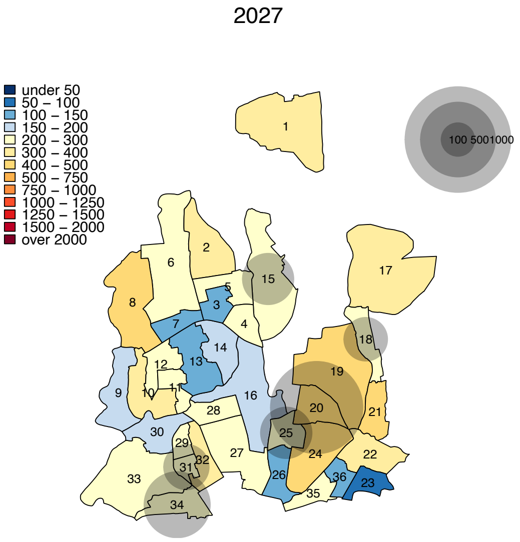
2027
under 50
50 – 100
100 – 150
150 – 200
200 – 300
300 – 400
400 – 500
500 – 750
750 – 1000
1000 – 1250
1250 – 1500
1500 – 2000
over 2000
100 5001000

(d)

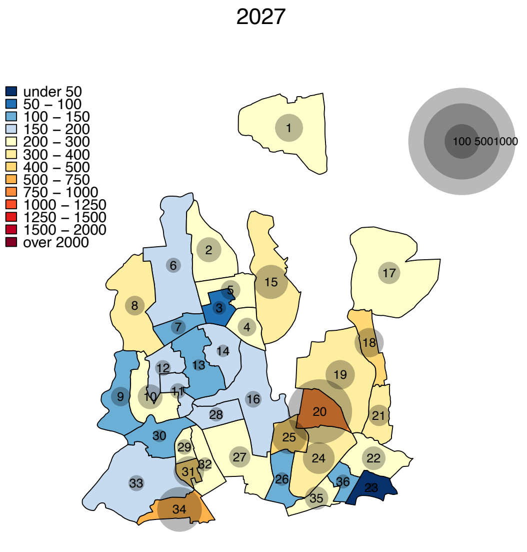
2027
under 50
50 – 100
100 – 150
150 – 200
200 – 300
300 – 400
400 – 500
500 – 750
750 – 1000
1000 – 1250
1250 – 1500
1500 – 2000
over 2000
100 5001000

(e)

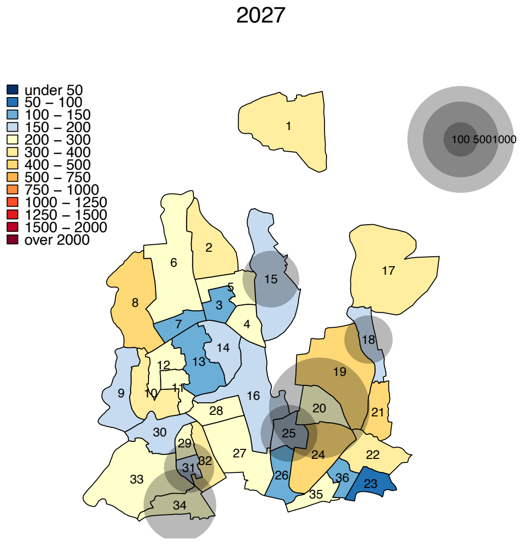
2027
under 50
50 – 100
100 – 150
150 – 200
200 – 300
300 – 400
400 – 500
500 – 750
750 – 1000
1000 – 1250
1250 – 1500
1500 – 2000
over 2000
100 5001000

(f)

### APPENDIX II. Multivariate Uncertainty analyses

We used multivariable sensitivity analysis to test how model outputs vary depending on the values of input parameters. This procedure also serves as an uncertainty analysis, by evaluating a reasonable range over which our primary outputs might vary. We varied each input parameter over a uniform distribution bounded by ±10% of the primary estimate (to ensure fair comparison between parameters). Within that range, parameter estimates were randomly chosen via Latin Hypercube Sampling (LHS). Parameters and their ranges are listed in the **Table A1**.

**Table A1. Input parameters and ranges used in sensitivity analyses**

| Parameters | Baseline estimates | Range of estimates (uniform distribution) |
|---|---|---|
| Mortality | 0.02 | 0.018 – 0.022 |
| TB Mortality | 0.15 | 0.135 – 0.165 |
| Reinfection rate | 0.50 | 0.45 – 0.55 |
| Diagnosis rate | 0.67 | 0.603 – 0.737 |
| Treatment failure rate | 0.10 | 0.09 – 0.11 |
| Reactivation rate | 0.015 | 0.00135 – 0.00165 |
| Rapid progression rate | 0.10 | 0.09 – 0.11 |
| Reduced time to diagnosis after ACF | 0.33 (1/3) | 0.30 – 0.36 |
| PT adherence rate | 0.80 | 0.72 – 0.88 |
| PT efficacy | 0.60 | 0.54 – 0.66 |

TB: tuberculosis, ACF: active case finding, PT: preventive therapy

We generated 10,000 LHS samples and conducted model simulations under each corresponding parameter set. Model outputs evaluated included: (1) the projected 10-year TB incidence without any intervention and (2) the ratio of projected reduction in 10-year TB incidence, comparing targeted interventions to the corresponding untargeted (citywide) interventions. A summary of model outputs under these simulations is provided in **Table A2**.

**Table A2. Mean and standard deviation of model outputs across 10,000 Latin Hypercube Samples in which input parameters were varied**

| | Dhaka South City Corporation | | Dhaka North City Corporation | |
|---|---|---|---|---|
| | **Mean** | **Standard deviation** | **Mean** | **Standard deviation** |
| 10-year projected TB incidence without any intervention (per 100,000) | 843 | 286 | 367 | 87 |
| Ratio of projected reduction in 10-year TB incidence, comparing targeted ACF to citywide ACF | 1.50 | 0.08 | 1.83 | 0.12 |
| Ratio of projected reduction in 10-year TB incidence, comparing targeted PT to citywide PT | 1.34 | 0.04 | 1.65 | 0.08 |
| Ratio of projected reduction in 10-year TB incidence, comparing combination of ACF and PT delivered in a targeted fashion to citywide | 1.05 | 0.03 | 1.32 | 0.05 |

TB: tuberculosis, ACF: active case finding, PT: preventive therapy

We computed partial rank correlation coefficients (PRCC) describing the correlation between each input parameter and each model output, adjusting for values of other input parameters. The PRCC ranked each parameter and output by magnitude and measured the sensitivity of an output variable to each parameter. PRCC values range from -1 (perfect negative correlation between input parameter and model output) to +1 (perfect positive correlation), with values of 0 indicating no correlation. The PRCC results were generated in Dhaka South City Corporation (DSCC) and Dhaka North City Corporation (DNCC) separately to account for different spatiotemporal patterns of TB incidence in the two regions. **[Figure A1—A3]**

**Figure A1. Partial rank correlation coefficient between input parameters and 10-year projected TB incidence (without any intervention).** Shown are partial rank correlation coefficients (PRCCs) describing the correlation between input parameter and the projected incidence in Dhaka South City Corporation (DSCC, panel a) and Dhaka North City Corporation (DNCC, panel b). PRCCs are calculated across 10,000 simulations in which parameter values were drawn using Latin Hypercube Sampling. Bars to the left (approaching -1.0) indicate negative correlations, and bars to the right (approaching 1.0) indicate positive correlations. The reactivation rate refers to the annual rate of reactivation from latent to active TB. The diagnosis rate refers to the annual rate at which populations with active TB are diagnosed and treated. Treatment failure denotes a proportion of those treated who remain in the active TB state. Relative protection against reinfection refers to the relative rate of reinfection among populations already latently infected, compared to the rate of initial infection among populations who are uninfected.

a. **Dhaka South City Corporation**

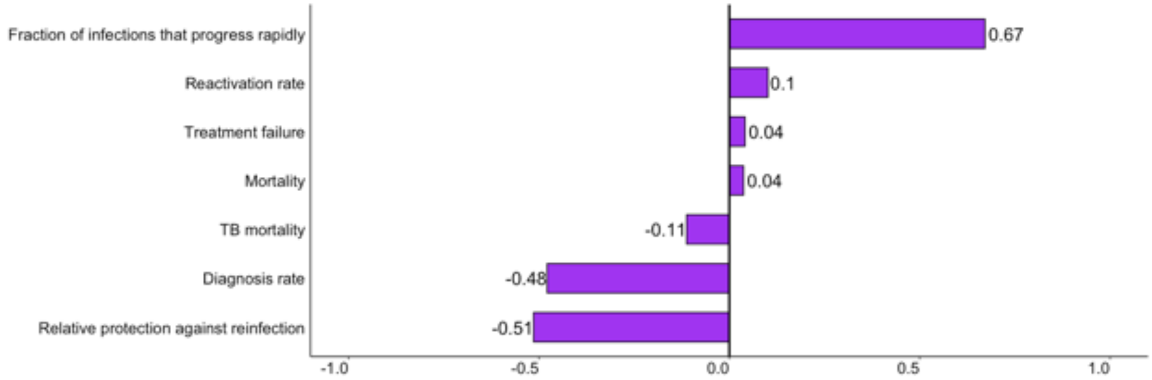

b. **Dhaka North City Corporation**

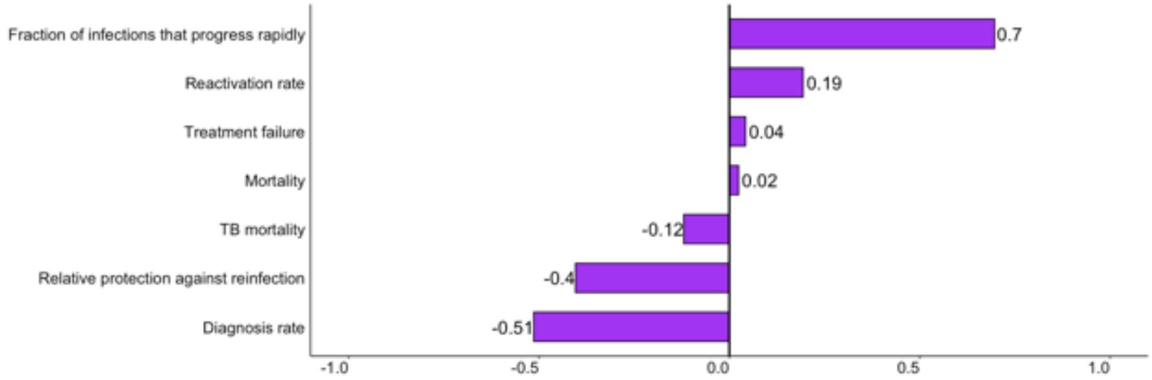

As expected, increasing the fraction of TB infection that progresses rapidly (a proxy for the transmission rate) and the reactivation rate resulted in increases in projected TB incidence, whereas increasing the rate of TB diagnosis, assumed protection against reinfection among people with latent TB infection, and mortality all resulted in declines in projected TB incidence. The strongest determinants of projected 10-year TB incidence in both DNCC and DSCC (Fig A1) were assumptions regarding TB transmission (i.e., number of infections that progress to TB disease) and TB diagnosis and treatment (i.e., the diagnosis rate).

**Figure A2. Partial rank correlation coefficient between model parameters and the relative reduction in 10-year TB incidence, comparing targeted to untargeted (citywide) interventions in Dhaka South City Corporation.** Shown are partial rank correlation coefficients (PRCCs) describing the correlation between input parameters and the relative reduction in 10-year TB incidence, comparing targeted to untargeted interventions consisting of (a) active case finding (ACF); (b) preventive therapy (PT); and (c) both. Bars to the left (approaching -1.0) indicate negative correlations, and bars to the right (approaching 1.0) indicate positive correlations. The reactivation rate refers to the annual rate of reactivation from latent to active TB. The diagnosis rate refers to the annual rate at which populations with active TB are diagnosed and treated. Treatment failure denotes a proportion of those treated who remain in the active TB state. Relative protection against reinfection refers to the relative rate of reinfection among populations already latently infected, compared to the rate of initial infection among populations who are uninfected. Reduction in time to diagnosis after ACF is tested under the assumption that the ACF implementation would reduce transmission by finding cases earlier. The PT efficacy and adherence rate are tested under the assumption that the PT implementation would reduce reactivation and rapid progression of infections.

**(a) After active case finding**

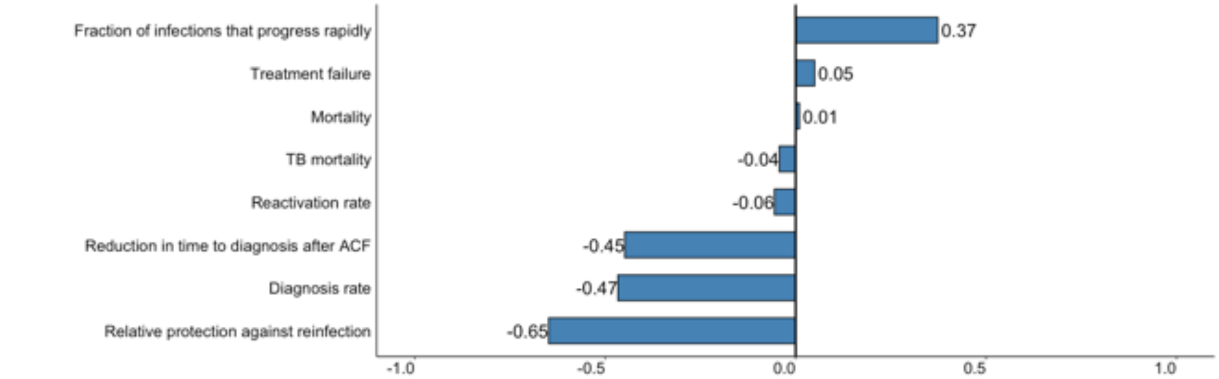

**(b) After Preventive therapy**

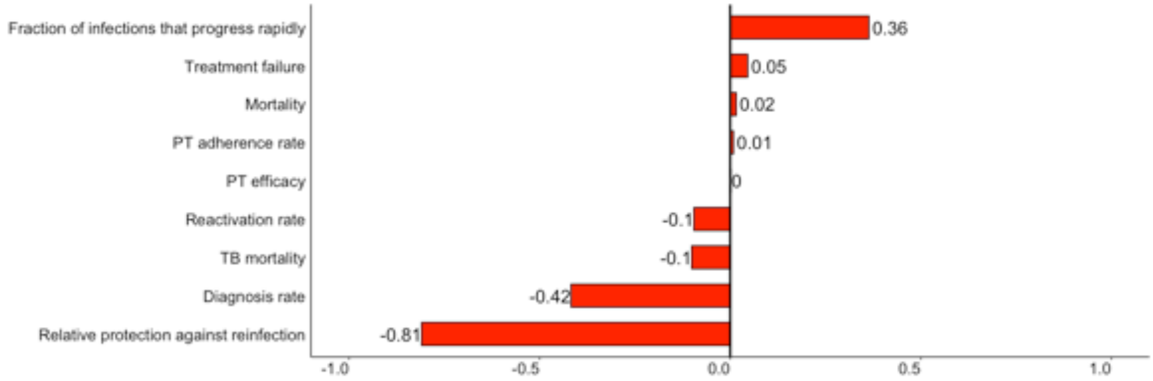

**(c) After both interventions**

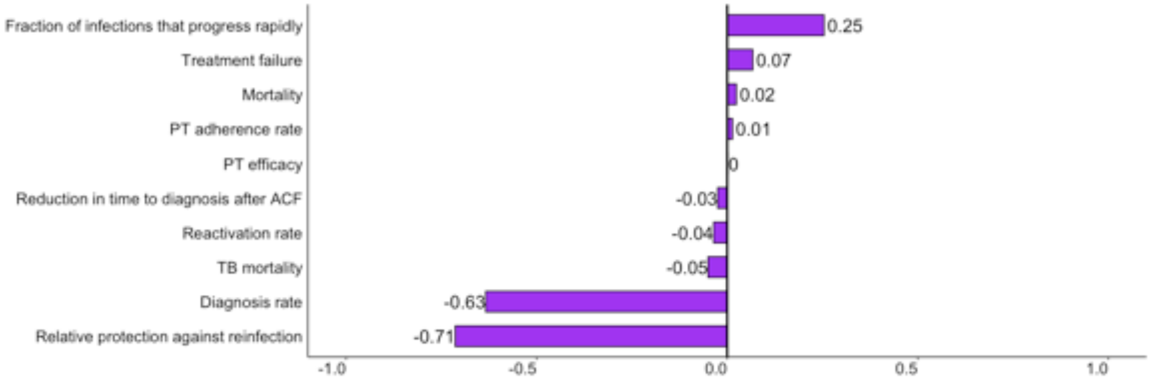

**Figure A3. Partial rank correlation coefficient between input parameters and the relative reduction in 10-year TB incidence, comparing targeted to untargeted (citywide) interventions in Dhaka North City Corporation.** Shown in bars are partial correlation coefficients (PRCCs) describing the correlation between input parameter values (as ranks) and the relative reduction in 10-year TB incidence, comparing targeted to untargeted interventions consisting of (a) active case finding; (b) preventive therapy; and (c) both. Bars to the left (approaching -1.0) indicate negative correlations, and bars to the right (approaching 1.0) indicate positive correlations. The reactivation rate refers to the annual rate of reactivation from latent to active TB. The diagnosis rate refers to the annual rate at which populations with active TB are diagnosed and treated. Treatment failure denotes a proportion of those treated who remain in the active TB state. Relative protection against reinfection refers to the relative rate of reinfection among populations already latently infected, compared to the rate of initial infection among populations who are uninfected. Reduction in time to diagnosis after ACF is tested under the assumption that the ACF implementation would reduce transmission by finding cases earlier. The IPT efficacy and adherence rate are tested under the assumption that the IPT implementation would reduce reactivation and rapid progression of infections.

#### (a) After active case finding

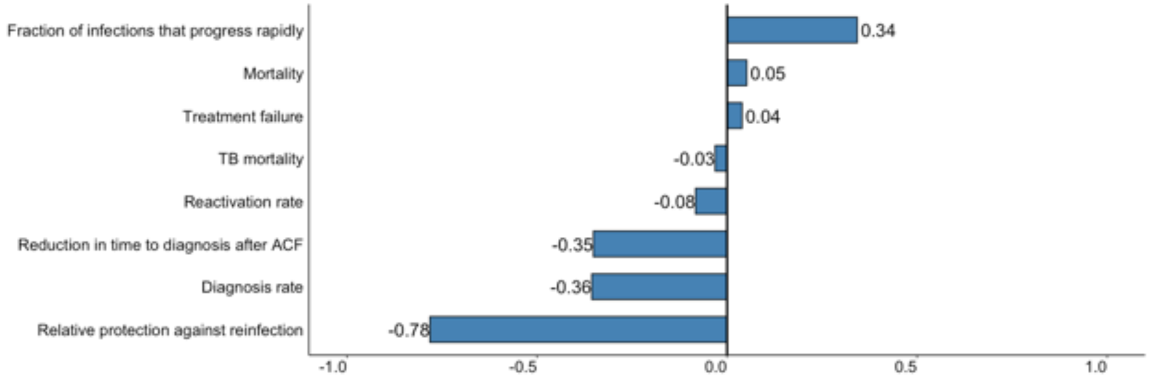

#### (b) After preventive therapy

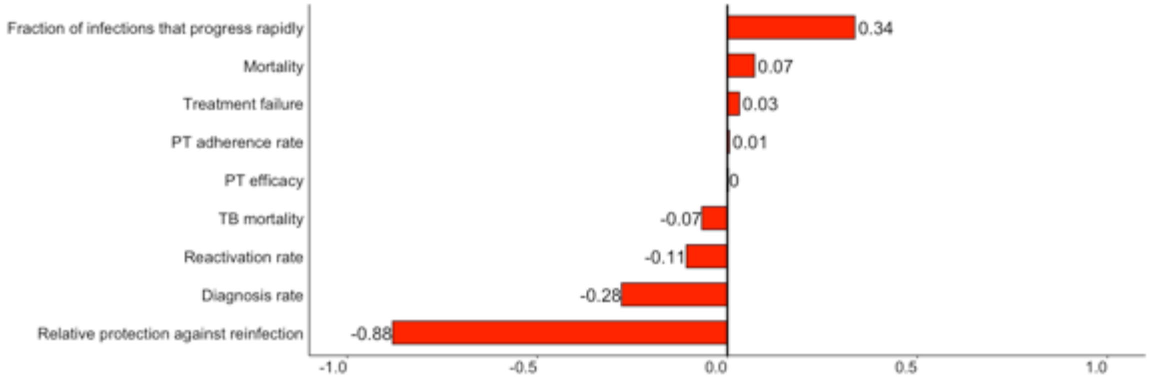

#### (c) After both interventions

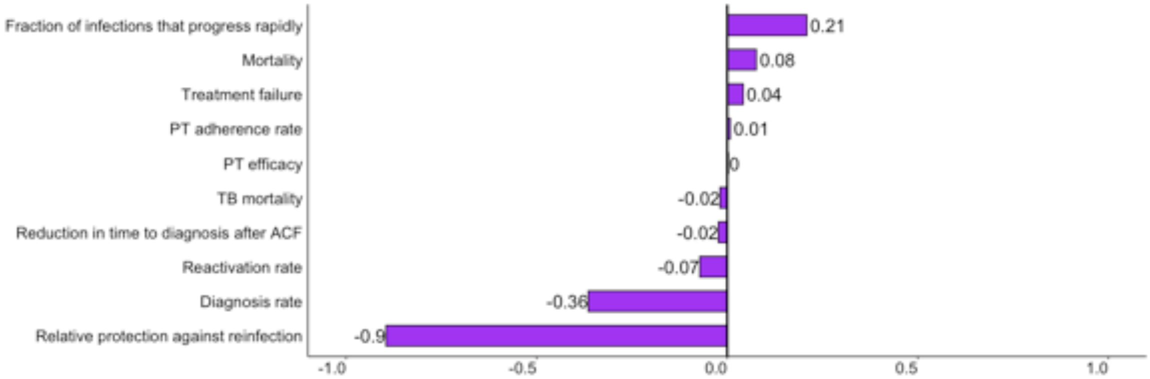

In both DSCC and DNCC, the strongest determinants of the relative value of targeting interventions were the assumed level of protection against reinfection and the rate of TB diagnosis (Figs S2 and S3). In settings where latent TB infection is assumed to provide strong protection against reinfection, the relative value of targeted interventions is reduced because a greater fraction of incident TB results from pre-existing infections (or reactivations) than from recent infection events (which are more amenable to targeted intervention). Similarly, when the underlying TB diagnosis and treatment rate is high, targeted interventions have less incremental value because each case of TB generates fewer secondary transmissions – again resulting in a scenario where the majority of incident TB reflects older rather than newer infection events. These dynamics (i.e., targeted interventions have greatest impact in settings where more incident TB is due to recent rather than remote infection) hold regardless of whether the intervention is targeted toward case finding or preventive therapy (PT). The one parameter that influences the value of targeted ACF but not of targeted PT is the estimated reduction in time to diagnosis after ACF – in settings where ACF is more effective in reducing time to diagnosis, the effect of targeting is greater. That this parameter does not strongly influence the combined intervention suggests that, when ACF and PT are combined, active cases would be prevented by PT, and hence the value of targeting is more strongly driven by PT than by ACF in our simple model.
